# Who is at risk of poor mental health following COVID-19 outpatient management?

**DOI:** 10.1101/2021.09.22.21263949

**Authors:** Katharina Hüfner, Piotr Tymoszuk, Dietmar Ausserhofer, Sabina Sahanic, Alex Pizzini, Verena Rass, Matyas Galffy, Anna Böhm, Katharina Kurz, Thomas Sonnweber, Ivan Tancevski, Stefan Kiechl, Andreas Huber, Barbara Plagg, Christian Wiedermann, Rosa Bellmann-Weiler, Herbert Bachler, Günter Weiss, Giuliano Piccoliori, Raimund Helbok, Judith Löffler-Ragg, Barbara Sperner-Unterweger

**Affiliations:** Department of Psychiatry, Psychotherapy and Psychosomatics, University Hospital for Psychiatry II, Medical University of Innsbruck, Innsbruck, Austria; Data Analytics As a Service Tirol, Innsbruck, Austria; Department of Internal Medicine II, Medical University of Innsbruck, Innsbruck, Austria; Institute of General Practice and Public Health, Claudiana Bolzano, Italy; Department of Neurology, Medical University of Innsbruck, Innsbruck, Austria; Tyrolean Federal Institute for Integrated Care, Innsbruck, Austria; Institute of General Medicine, Medical University of Innsbruck, Innsbruck, Austria

**Keywords:** COVID-19, SARS-CoV-2, mental health, depression, anxiety, mental stress, neurocognitive, long-COVID, post-COVID-19 syndrome

## Abstract

**Background:** COVID-19 convalescents are at risk of developing a de novo mental health disorder or of worsening of a pre-existing one. The objectives of our study was to phenotype individuals at highest risk of mental health disorders among COVID-19 outpatients.

**Methods:** We conducted a binational online survey study with adult non-hospitalized COVID-19 convalescents (Austria/AT: n=1157, Italy/IT: n= 893). Primary endpoints were positive screening for depression and anxiety (PHQ-4, Patient Health Questionnaire) and self-perceived overall mental health and quality of life rated with 4 point Likert scales. Psychosocial stress was surveyed with a modified PHQ stress module. Associations of the mental health with socio-demographic variables, COVID-19 course and recovery data were assessed by multi-parameter random forest and serial univariable modeling. Mental disorder risk subsets were defined by self-organizing map and hierarchical clustering algorithms. The survey analyses are publicly available (https://im2-ibk.shinyapps.io/mental_health_dashboard/).

**Results:** In the study cohorts, 4.6 (IT)/6% (AT) of participants reported depression and/or anxiety before to infection. At a median of 79 days (AT)/96 days (IT) post COVID-19 onset, 12.4 (AT)/19.3% (IT) of subjects were screened positive for anxiety and 17.3 (AT)/23.2% (IT) for depression. Over one-fifth of the respondents rated their overall mental health (AT: 21.8%, IT: 24.1%) or quality of life (AT: 20.3%, IT: 25.9%) as fair or poor. In both study collectives, psychosocial stress, high numbers of acute and persistent COVID-19 complaints and the presence of acute neurocognitive symptoms (impaired concentration, confusion, forgetfulness) were the strongest correlates of deteriorating mental health and poor quality of life. In clustering analysis, these variables defined a ‘high risk’ subset with particularly high propensity of post-COVID-19 mental health impairment and decreased quality of life. Pre-existing depression or anxiety was associated with an increased symptom burden during acute COVID-19 and recovery.

**Conclusion:** Our study revealed a bidirectional relationship between COVID-19 symptoms and mental health. We put forward specific acute symptoms of the disease as ‘red flags’ of mental health deterioration which should prompt general practitioners to identify COVID-19 patients who may benefit from early psychological and psychiatric intervention.

**Trial registration:** ClinicalTrials.gov: NCT04661462.

## Background

Prevalence of mental health disorders rose during the COVID-19 pandemic in the general population from 4% in 2006(1) to 20% for depression and from 5% in 2008(2) to 19% for anxiety as of March 2020(3). The mental health deterioration following COVID-19 was described primarily for hospitalized subjects(4). The frequency of depression or anxiety in inpatients was estimated for approximately 25% at 5 – 12 months post infection (5–7). A real-world analysis of 62,354 COVID-19 in- and outpatients at 14 – 90 days follow-up revealed the overall incidence of psychiatric conditions at 18.1% [95%CI: 17.6 to 18.6], out of which 5.8% [5.2–6.4] comprised de novo disorders(8). In this study, a pre-existing mental illness was put forward as a risk factor for SARS-CoV-2 infection suggestive of a bi-directional relationship between psychiatric conditions and COVID-19(8). A large, medical record-based comparison of long-term sequelae in COVID-19 and non-COVID-19 healthcare system users revealed an excess of sleep/wake-(relative risk: 14.5 [11.5–17.3]), anxiety/fear-(5.4 [3.4–7.3]) and trauma/stress-related disorders (8.9 [6.6–11.1]) in COVID-19 patients(9). Still, the prevalence and risk factors of mental health conditions and diminished quality of life in COVID-19 outpatients, which may be missed from medical record analyses, are insufficiently characterized.

The binational ‘Health after COVID-19 in Tyrol’ study aims at exploring the disease course as well as physical and mental recovery in two cohorts of non-hospitalized convalescents(10). Herein, using multi-parameter modeling we sought to assess the impact of demographics, socioeconomics, comorbidities, COVID-19 disease symptoms and course and the psychosocial stress on the anxiety, depression, self-perceived overall mental health and quality of life. By association analysis, we aimed to identify individuals at risk of worsening mental health and quality of life, which may particularly benefit from early psychological and psychiatric support. Finally, we made the study results publicly available as an online dashboard (https://im2-ibk.shinyapps.io/mental_health_dashboard/)(11).

## Methods

### Study design and approval

The multi-center online survey study ‘Health after COVID-19 in Tyrol’ (ClinicalTrials.gov: NCT04661462) was conducted between the 30^th^ September 2020 and 11^th^ July 2021 in two cohorts independently recruited in Tyrol/Austria (AT) and South Tyrol/Italy (IT)(10). The study inclusion criteria were residency in the study regions, age of ≥16 (AT) or ≥18 years (IT) and a laboratory-confirmed SARS-CoV-2 infection (PCR or seropositivity). The respondents with a minimum observation time of < 28 days between the infection diagnosis and survey completion or hospitalized because of COVID-19 were excluded from the analysis (**Figure 1**). The participants were invited by a public media call (AT and IT) or by their general practitioners (IT).

**Figure 1.**
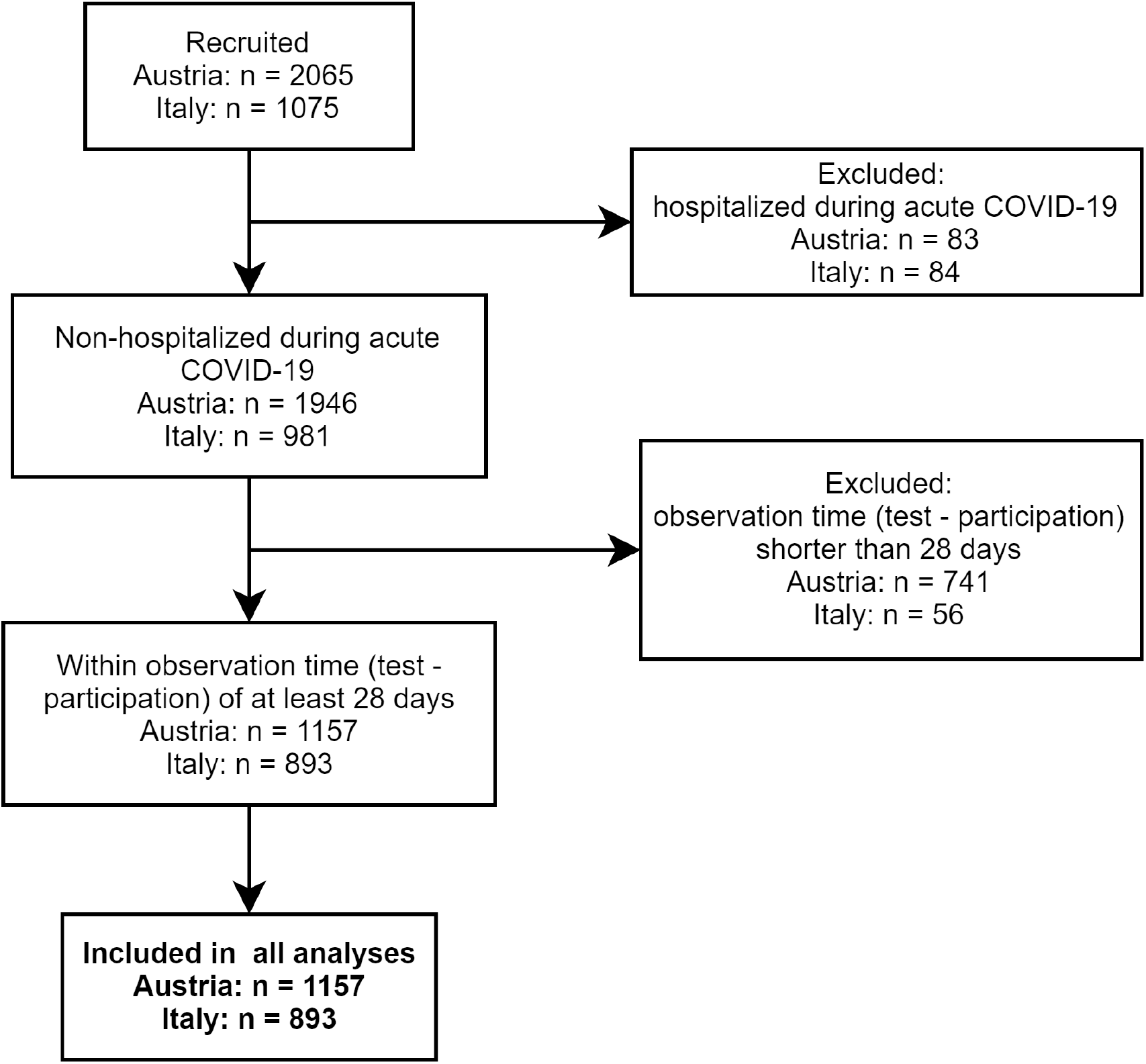
CONSORT flow diagram for the study cohorts.

The study was conducted in accordance with the Declaration of Helsinki and the European data policy. Each participant gave a digitally signed informed consent to participate. The study protocol was approved by the institutional review boards of the Medical University of Innsbruck (AT, approval number: 1257/2020) and of the Autonomous Province of Bolzano – South Tyrol (IT: 0150701).

### Measures, definitions and data transformation

The detailed description of the questionnaire variables is provided by Sahanic et al.(10), in **Supplementary Methods** and **Supplementary Table S1**.

Symptoms were classified as acute complaints present during the first 2 weeks after clinical onset and persistent symptoms present for ≥ 4 weeks (10,12). Confusion, impaired concentration and forgetfulness were subsumed under ‘neurocognitive symptoms’.

Pre-existing depression/anxiety or sleep disorders were surveyed as single question each (yes/no). Self-perceived overall mental health (OMH) and quality of life (QoL) were rated with a 4-point Likert scale (‘excellent’, ‘good’, ‘fair’, ‘poor’, scored: 0, 1, 2, 3). Anxiety and depression at the survey completion were investigated using PHQ-4 questionnaire(11,13), with ≥ 3 point cutoffs for the clinical signs of depression (DPR) or anxiety (ANX). Psychosocial stress was measured with a modified 7 item (answers: ‘no’, ‘little’, ‘some’, ‘a lot’, scored: 0, 1, 2, 3) PHQ stress module(11,14), without items on weight, sexuality and past traumatic/serious events; the item on worries/dreams was adapted to COVID-19. The stress scoring was re-coded as quartile strata encompassing 0 – 2, 3 – 4, 5 – 6, 7 – 21 points. Substantial psychosocial stress was defined by a ≥ 7 point cutoff.

### Statistical analysis

Data were analyzed with R version 4.0.5 (15,16). Statistical significance of variable median or distribution differences between groups was determined by U, Kruskal-Wallis or χ^2^ test, as appropriate. Categorical variable co-occurrence was assessed by Cohen’s κ and Z test. P values were corrected for multiple comparisons with Benjamini-Hochberg method(17).

Random forest models for ANX, DPR, OMH and QoL scoring were built and cross-validated (10 fold) separately in the AT and IT cohort(18,19). To account for possible effects of diagnosis – survey time, a stratified observation time variable was included in the models (**Supplementary Table S1**). The factor’s influence on the scoring variable was measured by the difference in model’s mean squared error (ΔMSE) as described in (18). To assess the factor impact on the combined mental health and quality of life rating, centered principal component analysis (PCA) with factor’s normalized values of ΔMSE for the OMH, QoL, ANX and DPR scoring was performed(20). The factors with the 10 largest PCA loadings in each cohort were used further for univariable modeling and clustering. Univariable modeling was accomplished with age- and sex-weighted Poisson regression(10). Clustering was done with the self-organized map procedure (SOM, 11×11 unit hexagonal grid, Jaccard distance) and subsequent hierarchical clustering (Ward D2 algorithm, Euclidean distance)(21,22). Details of statistical analysis are provided in **Supplementary Methods**.

## Results

### Sociodemographics and clinical characteristics of the study cohorts

In total, 1157 questionnaires in the AT and 893 in the IT cohort were analyzed (**Figure 1**). Detailed characteristics of the cohorts were reported by Sahanic et al. (10). In brief, study participants were predominantly working-age (31 – 65 years: AT: 71.9%, IT: 77.8%), female (AT: 65.1%, IT: 68.3%) and actively employed (> 80%). Pre-existing co-morbidities were declared by 41.2 (IT)/49.7% (AT). Depression or anxiety (AT: 6%, IT: 4.6%) and sleep disorders (AT: 4.6%, IT: 4%) before COVID-19 were reported by roughly 1 of 20 respondents (**Table 1**). Notably, the overlap between the pre-existing depression/anxiety and sleep problems was only minute (AT: Cohen’s κ = 0.21 [95%CI: 0.1 – 0.31], IT: κ = 0.17 [0.048 – 0.3]). The collectives significantly differed in language, education and employment structure and the time interval between the diagnosis and survey completion (**Table 1, Supplementary Table S2**).

**Table 1.**
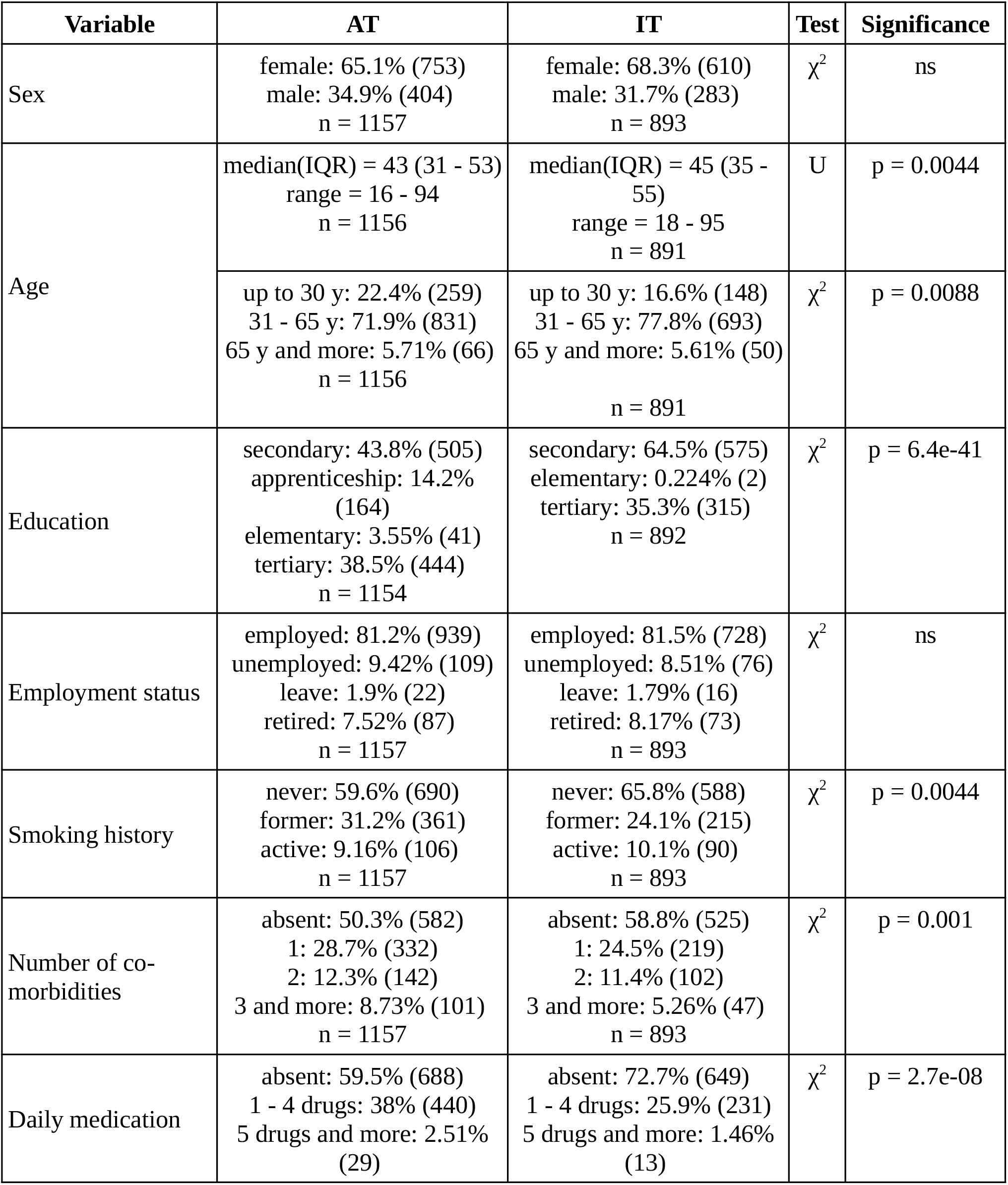
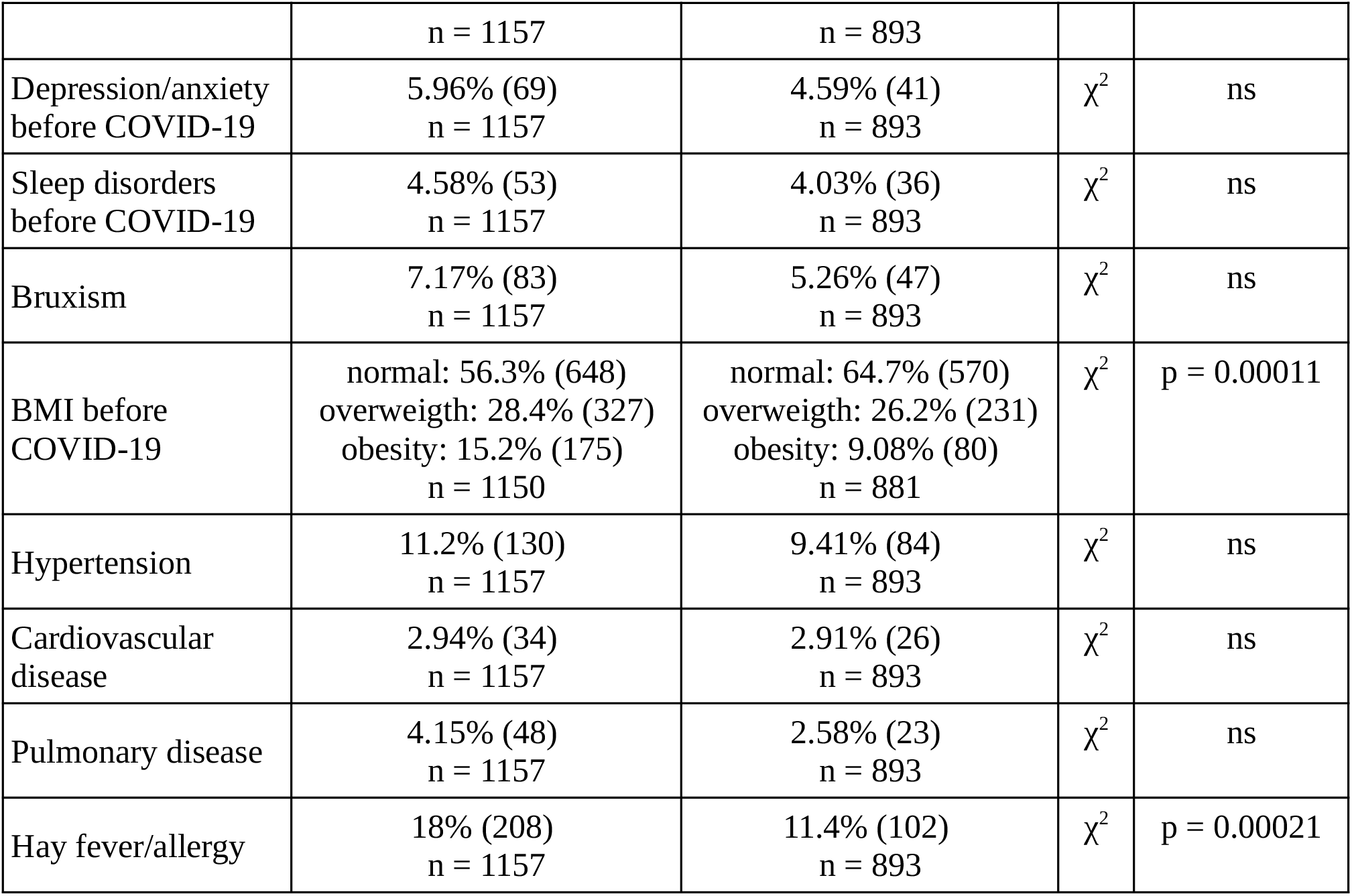
Baseline characteristic of the study cohorts. AT: Austria/Tyrol cohort, IT: Italy/South Tyrol cohort, Test: statistical test used for the AT vs IT comparison, Significance: test p value corrected for multiple comparisons with Benjamini-Hochberg method.

The percentage of asymptomatic cases ranged between 8.3 (AT) and 12.3% (IT) (**Table 2, Supplementary Table S2**). Respondents declared a median of 13 complaints (out of 44 features queried, IQR: AT: 9 – 18, IT: 7 – 18) present in the first two weeks after clinical onset. Persistent symptoms lasting for ≥28 days(10,12) were discerned in 47.6 (AT)/49.3% (IT). Roughly half of the participants suffered from acute neurocognitive symptoms (AT: 48%, IT: 50.4%) such as memory or concentration deficits or confusion, in 18.2 (AT)/22.6% (IT) at least one persistent neurocognitive symptom was present (**Table 2**).

**Table 2.** Characteristic of the course of SARS-CoV2 infection and convalescence in the study cohorts. AT: Austria/Tyrol cohort, IT: Italy/South Tyrol cohort, Test: statistical test used for the AT vs IT comparison, Significance: test p value corrected for multiple comparisons with Benjamini-Hochberg method.

| Variable | AT | IT | Test | Significance |
| --- | --- | --- | --- | --- |
| Acute COVID-19 symptoms | 91.7% (1060)<br>n = 1156 | 87.7% (782)<br>n = 892 | $\chi^2$ | p = 0.0068 |
| Number of acute symptoms | median(IQR) = 13 (9 - 18)<br>range = 0 - 42<br>n = 1156 | median(IQR) = 13 (7 - 18)<br>range = 0 - 39<br>n = 892 | U | ns |
| Number of acute neurocognitive symptoms | median(IQR) = 1 (0 - 2)<br>range = 0 - 3<br>n = 1157 | median(IQR) = 0 (0 - 2)<br>range = 0 - 3<br>n = 893 | U | ns |
| | 0: 49.6% (574)<br>1: 20.4% (236)<br>2: 17% (197)<br>3: 13% (150)<br>n = 1157 | 0: 52% (464)<br>1: 14.2% (127)<br>2: 16.3% (146)<br>3: 17.5% (156)<br>n = 893 | $\chi^2$ | p = 0.0015 |
| Persistent COVID-19 symptoms | 47.6% (550)<br>n = 1156 | 49.3% (440)<br>n = 892 | $\chi^2$ | ns |
| Number of persistent symptoms | median(IQR) = 0 (0 - 3)<br>range = 0 - 34<br>n = 1156 | median(IQR) = 0 (0 - 3)<br>range = 0 - 29<br>n = 892 | U | ns |
| Number of persistent neurocognitive symptoms | median(IQR) = 0 (0 - 0)<br>range = 0 - 3<br>n = 1157 | median(IQR) = 0 (0 - 0)<br>range = 0 - 3<br>n = 893 | U | p = 0.0068 |
| | 0: 81.8% (946)<br>1: 7.26% (84)<br>2: 7.78% (90)<br>3: 3.2% (37)<br>n = 1157 | 0: 77.4% (691)<br>1: 5.6% (50)<br>2: 9.63% (86)<br>3: 7.39% (66)<br>n = 893 | $\chi^2$ | p = 0.00032 |

At the time of study completion, i. e. approximately 12 weeks post clinical COVID-19 onset (AT, median: 79 days [IQR: 40 – 175], IT: 96 [60 – 138]), over one-fifth of the participants rated their overall mental health (AT: 21.8%, IT: 24.1%) or quality of life (AT: 20.3%, IT: 25.9%) as fair or poor (**Table 3, Supplementary Table S2** and (11). At this time point, anxiety (ANX) was observed in 12.4 (AT)/19.3% (IT), signs of depression (DPR) in 17.3 (AT)/23.2% (IT) and substantial psychosocial stress in 21.3 (AT)/25.6% (IT) of the respondents. Except for the poor/rair rating of self-perceived OMH and QoL, the studied mental status features demonstrated only weak co-occurrence (Cohen’s κ < 0.5, **Supplementary Figure S1**). Importantly, the investigated mental health and quality of life rating variables were weakly associated with the observation time (**Supplementary Figure S2** and (11)). The QoL rating as well as prevalence of ANX, DPR and substantial stress were significantly higher in the IT than in the AT study collective (**Table 3** and (11)).

**Table 3.** Rating of the mental health following COVID-19 in the study cohorts. AT: Austria/Tyrol cohort, IT: Italy/South Tyrol cohort, Test: statistical test used for the AT vs IT comparison, Significance: test p value corrected for multiple comparisons with Benjamini-Hochberg method.

| Variable | AT | IT | Test | Significance |
| --- | --- | --- | --- | --- |
| Overall Mental Health | poor: 3.46% (40)<br>fair: 18.3% (212)<br>good: 48.6% (562)<br>excellent: 29.6% (343)<br>n = 1157 | poor: 2.91% (26)<br>fair: 21.2% (189)<br>good: 48.2% (430)<br>excellent: 27.8% (248)<br>n = 893 | $\chi^2$ | ns |
| Overall Mental Health Score | mean(SD) = 0.956 (0.785)<br>median(IQR) = 1 (0 - 1)<br>range = 0 - 3<br>n = 1157 | mean(SD) = 0.992 (0.779)<br>median(IQR) = 1 (0 - 1)<br>range = 0 - 3<br>n = 893 | U | ns |
| Quality of Life | poor: 4.32% (50)<br>fair: 16% (185)<br>good: 51% (590)<br>excellent: 28.7% (332)<br>n = 1157 | poor: 3.36% (30)<br>fair: 22.5% (201)<br>good: 54.3% (485)<br>excellent: 19.8% (177)<br>n = 893 | $\chi^2$ | p = 8.3e-06 |
| Quality of Life Score | mean(SD) = 0.959 (0.787)<br>median(IQR) = 1 (0 - 1)<br>range = 0 - 3<br>n = 1157 | mean(SD) = 1.09 (0.741)<br>median(IQR) = 1 (1 - 2)<br>range = 0 - 3<br>n = 893 | U | p = 2.6e-05 |
| DPR score | mean(SD) = 1.39 (1.58)<br>median(IQR) = 1 (0 - 2)<br>range = 0 - 6<br>n = 1154 | mean(SD) = 1.61 (1.68)<br>median(IQR) = 1 (0 - 2)<br>range = 0 - 6<br>n = 892 | U | p = 0.0073 |
| Depression Screening-positive | 17.3% (200)<br>n = 1154 | 23.2% (207)<br>n = 892 | $\chi^2$ | p = 0.0023 |
| Anxiety score | mean(SD) = 0.949 (1.33)<br>median(IQR) = 0 (0 - 2)<br>range = 0 - 6<br>n = 1151 | mean(SD) = 1.35 (1.57)<br>median(IQR) = 1 (0 - 2)<br>range = 0 - 6<br>n = 893 | U | p = 4.1e-09 |
| Anxiety Screening-positive | 12.4% (143)<br>n = 1151 | 19.3% (172)<br>n = 893 | $\chi^2$ | p = 7.6e-05 |
| Psychosocial Stress Score | mean(SD) = 4.28 (3.53)<br>median(IQR) = 4 (2 - 6)<br>range = 0 - 19<br>n = 1153 | mean(SD) = 4.4 (3.54)<br>median(IQR) = 4 (2 - 7)<br>range = 0 - 19<br>n = 890 | U | ns |
| Substantial | 21.3% (246) | 25.6% (228) | $\chi^2$ | p = 0.035 |
| psychosocial stress | n = 1153 | n = 890 |  |  |

### Key factors impacting combined mental health and quality of life outcome in COVID-19 convalescents

To identify the crucial factors impacting the ANX, DPR, OMH and QoL scoring, we applied the random forest machine learning modeling(18) to a set of 145 explanatory variables in the AT and IT cohorts (**Supplementary Table S1**). Such models demonstrated good performances in the training data sets (Spearman’s ρ > 0.9, means absolute error (MAE): 0.21 – 0.44) and 10-fold cross-validation (MAE: 0.51 – 1.1) (**Supplementary Figures S3 – S6)**.

The high and moderate psychosocial stress levels (>75^th^ and >50^th^ percentile) affected the ANX, OMH and QoL rating to the greatest extent in each cohort (**Supplementary Figures S3, S5 – S6**). In turn, high psychosocial stress values along with acute concentration deficits and persistent tiredness displayed the tightest association with the DPR score (**Supplementary Figure S4**). Readouts of COVID-19 disease severity, duration and character, such as overall symptom numbers and the presence of neurocognitive symptoms were found among the 20 most influential factors for the ANX, DPR, OMH and QoL rating. Furthermore, self-reported mental health conditions before COVID-19 (depression/anxiety or sleep disorder) were significantly associated with poorer OMH, QoL, DPR and ANX scoring in both collectives (**Supplementary Figure S7** and (11)) and present among the most influential factors for the ANX, DPR, OMH and QoL scoring in at least one study cohort (**Supplementary Figures S3 – S6**).

To discern features influencing the combined mental health and quality of life scoring, principal component analysis was performed (20). This approach underscored the prime effect of moderate and psychosocial stress on the rating in both the AT and IT collective, followed by persistent tiredness and acute neurocognitive manifestations such as confusion, concentration and memory deficits as well as acute and persistent symptom burden (**Figure 2**).

**Figure 2.**
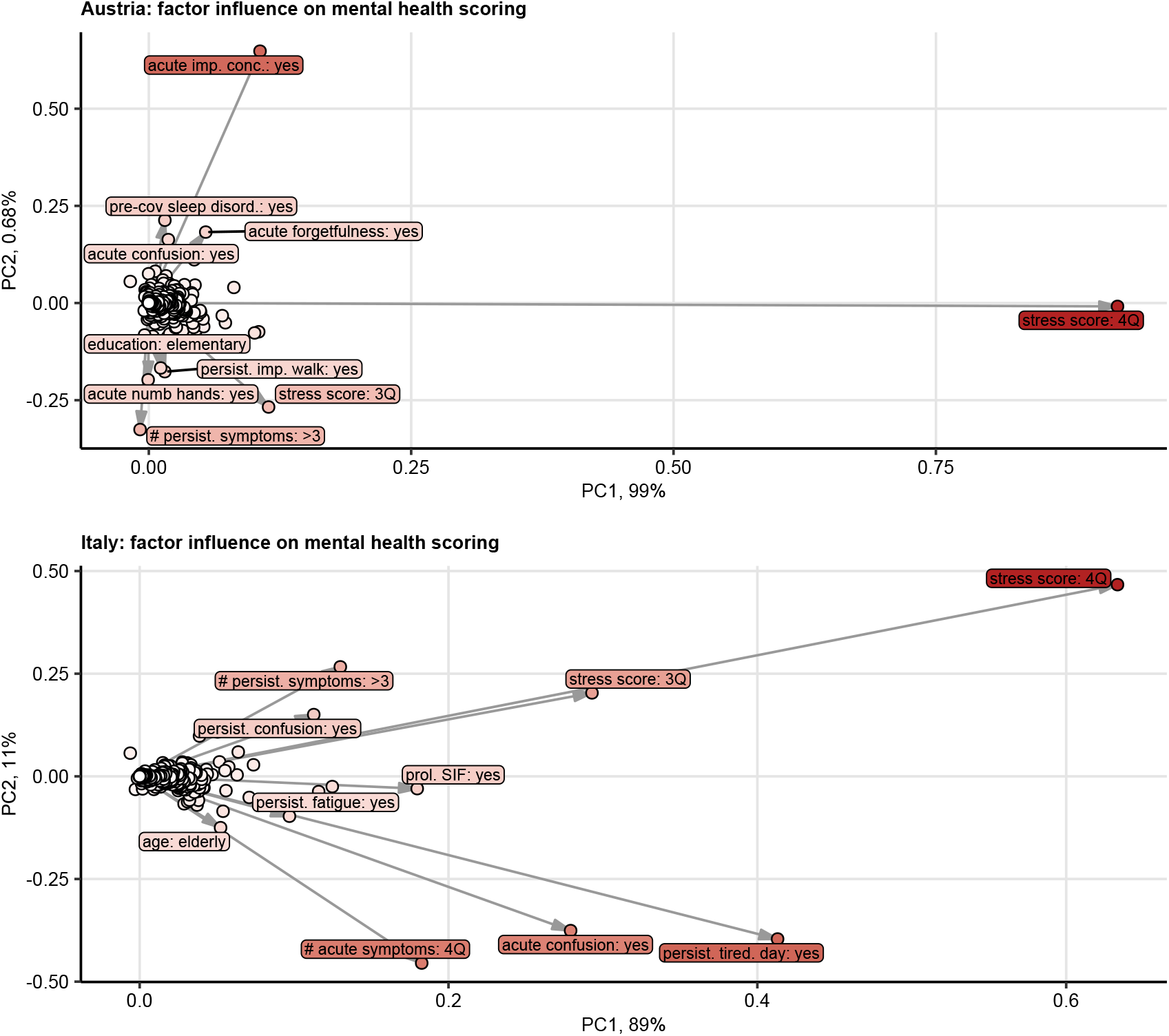
The most influential factors for the combined mental health and quality of life scoring following COVID-19. The effects of 145 demographic, clinical, socioeconomic and psychosocial factors (**Supplementary Table S1**) on the overall mental health, quality of life, anxiety and depression scoring were modeled with random forest technique (**Supplementary Figures S3 – S6**). The the impacts of candidate factors on the model fits were subjected to two-dimensional centered principal component analysis. Factors’ loadings in respect to the principal components (PC) are presented in the plots, percent variance is shown in the axes. Top 10 factors with the largest loadings vectors as a measure of the influence on the combined mental health and quality of life scoring were labeled, point color corresponds to the vector length. prol.: prolonged, SIF: severe illness feeling, imp.: impaired, conc.: concentration, #: number, tired.day.: tiredness at day, pre-cov sleep disord.: sleep disorder before COVID-19, 3Q, 4Q: 3rd and 4th quartile, persist.: persistent.

Importantly, by means of canonical age- and sex-weighted univariable Poisson regression, high psychosocial stress (> 75^th^ percentile) was found the strongest co-variate of poor ANX, DPR, OMH and QoL ratings each (OMH: pooled β = 2.74 [95%CI: 2.66 – 2.83], QoL: β = 2.48 [2.4 – 2.57], ANX: β = 6.04 [5.88 – 6.2], DPR: β = 4.34 [4.21 – 4.47](23)), followed by the high burden of acute infection symptoms, acute and persistent neurocognitive complaints (**Figure 3, Supplementary Tables S3 – S4**).

**Figure 3.**
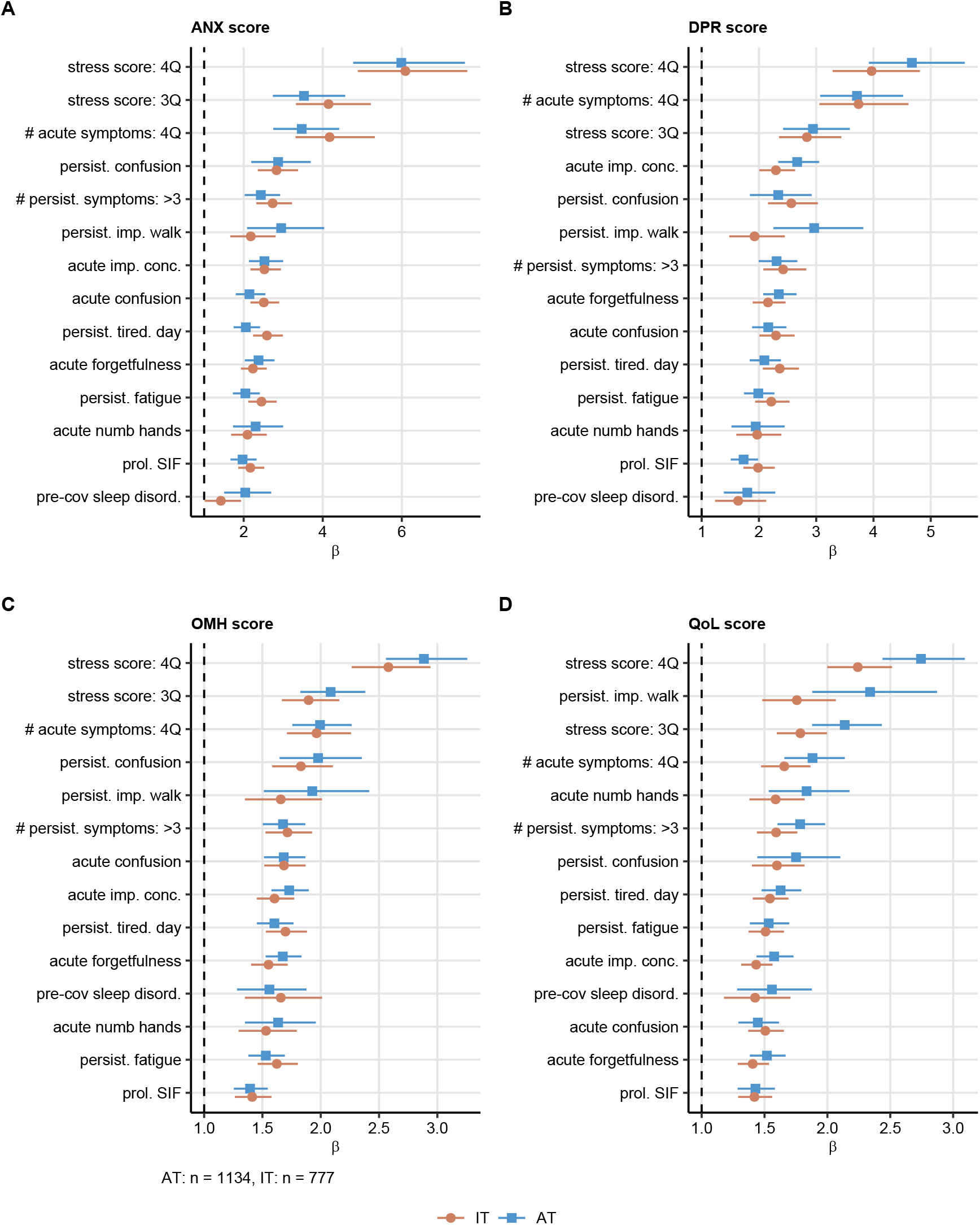
Correlation of the most influential factors with the mental health readouts investigated by univariable modeling. Correlation of the 10 most influential factors affecting the combined mental health and quality of life scoring (**Figure 2**) with the anxiety (ANX) (A), depression (DPR) (B), overall mental health (OMH) (C) and quality of life (QoL) (D) rating was investigated by univariable, age- and sex-weighted Poisson regression (**Supplementary Table S5**). β estimate values with 95% confidence intervals for the significant correlations in both the Austria/Tyrol (AT) and Italy/South Tyrol (IT) cohort are presented as forest plots. prol.: prolonged, SIF: severe illness feeling, imp.: impaired, conc.: concentration, #: number, tired.day.: tiredness at day, pre-cov sleep disord.: sleep disorder before COVID-19, 3Q, 4Q: 3rd and 4th quartile, persist.: persistent.

### Acute neurocognitive symptoms and poly-symptomatic acute COVID-19 define the subjects at risk of poor mental health

Next, we asked whether the set of the 10 most influential factors impacting the combined mental health and quality of life in the AT or IT cohort (**Figure 2**) may be applied to identify convalescents at particular risk of mental health deterioration following COVID-19.

By a self-organizing map and hierarchical clustering association analysis(21,22) three participants subsets termed ‘Low Risk’ (LR), ‘Intermediate Risk’ (IR) and ‘High Risk’ (HR) Mental Disorder Risk Clusters were discerned in each study cohort (**Figure 4, Supplementary Figure S8**). The prime hallmarks of the IR and HR subsets were highly frequent acute confusion, acute concentration and memory problems and highly poly-symptomatic acute COVID-19. The HR subset differed from the IR cluster by higher proportions of subjects with > 3 persistent symptoms, persistent tiredness and fatigue (**Figure 4, Supplementary Figure S9**).

**Figure 4.**
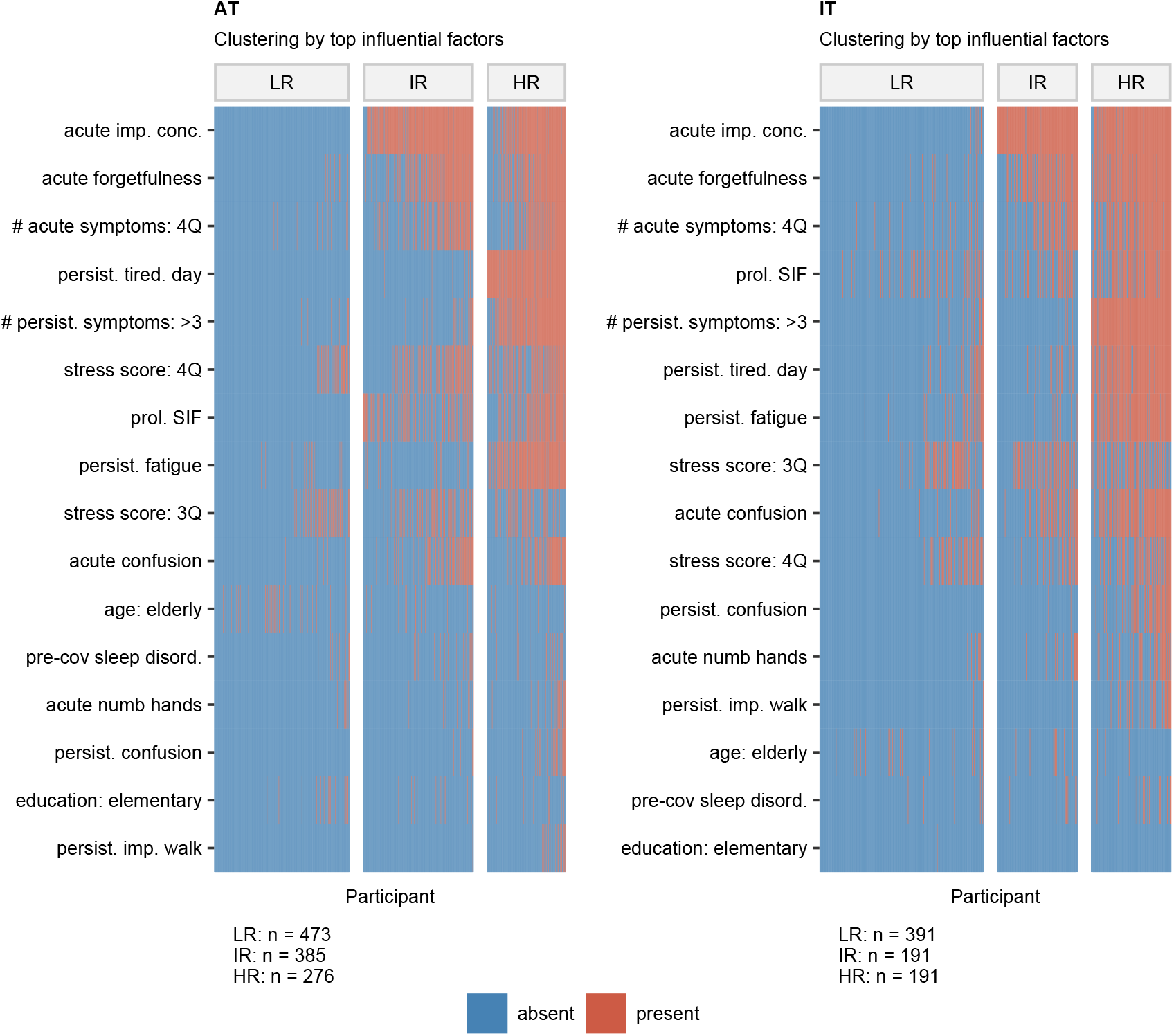
Clustering of the study participants by the most influential factors affecting the combined mental health and quality of life scoring. Study participants were assigned to the Low Risk (LR), Intermediate Risk (IR) and High Risk (HR) subsets by clustering analysis of the most influential factors affecting the combined mental health and quality of life scoring (**Figure 2**) by the self-organizing map (SOM, 11 × 11 hexagonal grid, Jaccard distance between participants) and the hierarchical clustering (Ward D2 method, Euclidean distance between the SOM nodes) algorithms. Presence/ absence of the features is presented as heat maps. N numbers of individuals assigned to the clusters are presented next to the plots. prol.: prolonged, SIF: severe illness feeling, imp.: impaired, conc.: concentration, #: number, tired.day.: tiredness at day, pre-cov sleep disord.: sleep disorder before COVID-19, 3Q, 4Q: 3rd and 4th quartile, persist.: persistent.

Notably, the HR followed by the IR group demonstrated significantly worse ANX, DPR, OMH and QoL rating as well as higher frequencies of anxiety (AT: 5.3% in LR, 24% in HR, IT: 9.7% in LR, 44% in HR) and depression (AT: 4.9% in LR, 34.8% in HR, IT: 11.3% in LR, 40.3% in HR) compared with the IR cluster (**Figure 5** and (11)). In addition, chest, abdomen and joint pain complaints, diminished appetite, sleep problems, dizziness and tachycardia during acute COVID-19 were significantly more frequent in the IR and HR cluster than in the LR subset (**Supplementary Figure S10, Supplementary Table S5**).

**Figure 5.**
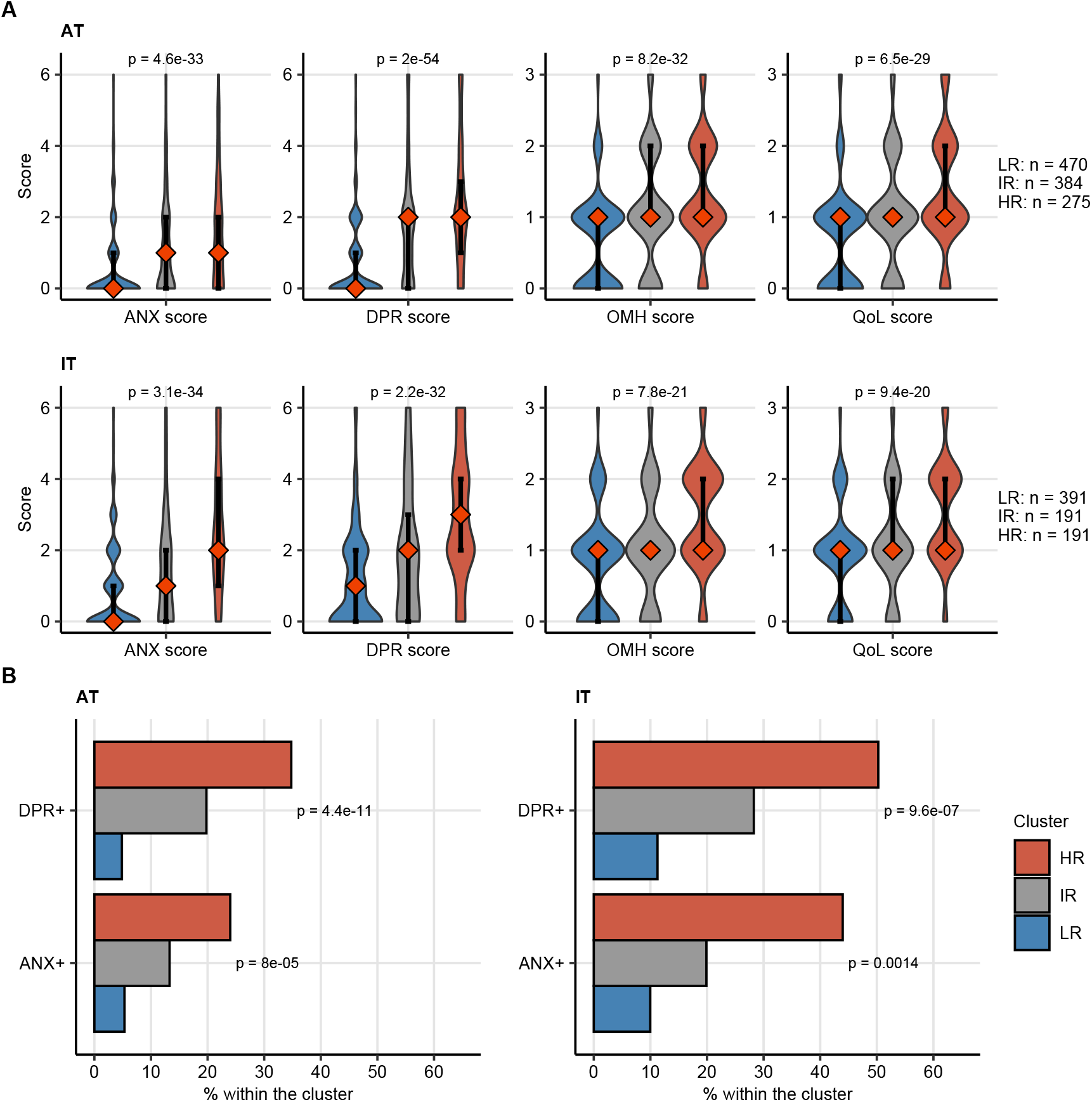
Mental health scoring, depression and anxiety prevalence in the mental disorder risk clusters. Study participants were assigned to the Low Risk (LR), Intermediate Risk (IR) and High Risk (HR) subsets as presented in **Figure 4**. (A) Rating of anxiety (ANX), depression (DPR), overall mental health (OMH) and quality of life (QoL) in the clusters presented as violin plots, diamonds with whiskers represent medians with IQRs. Statistical significance was assessed by Kruskal-Wallis test. (B) Frequency of positive depression (DPR+) and anxiety (ANX+) screening in the clusters. Statistical significance was assessed by χ^2^ test. P values corrected for multiple comparisons with Benjamini-Hochberg method are shown in the plots. N numbers of individuals assigned to the clusters are presented next to the plots.

### Depression or anxiety before COVID-19 is linked to a higher symptom burden and persistence

Finally, we sought to investigate whether a reciprocal link between the pre-existing psychiatric disorder and the burden of acute and persistent COVID-19 symptoms exists.

Participants declaring anxiety/depression before the infection had a 20% higher median burden of overall acute COVID-19 symptoms and >30% more acute neurocognitive symptoms compared with the depression/anxiety-free subset (**Figure 6A – C**). In addition, pre-existing depression/anxiety was linked to a significantly higher prevalence of long COVID both in the AT (depression/anxiety: 65.2% vs depression/anxiety-free: 46.4% long COVID, p = 0.0037, χ^2^ test) and IT cohort (68.3% vs 48.4% long COVID, p = 0.020, χ^2^ test). Accordingly, significantly higher counts of persistent overall and neurocognitive symptoms were observed in the pre-COVID-19 depression/anxiety subset compared with the depression/anxiety-free subjects (**Figure 6D – F**).

**Figure 6.**
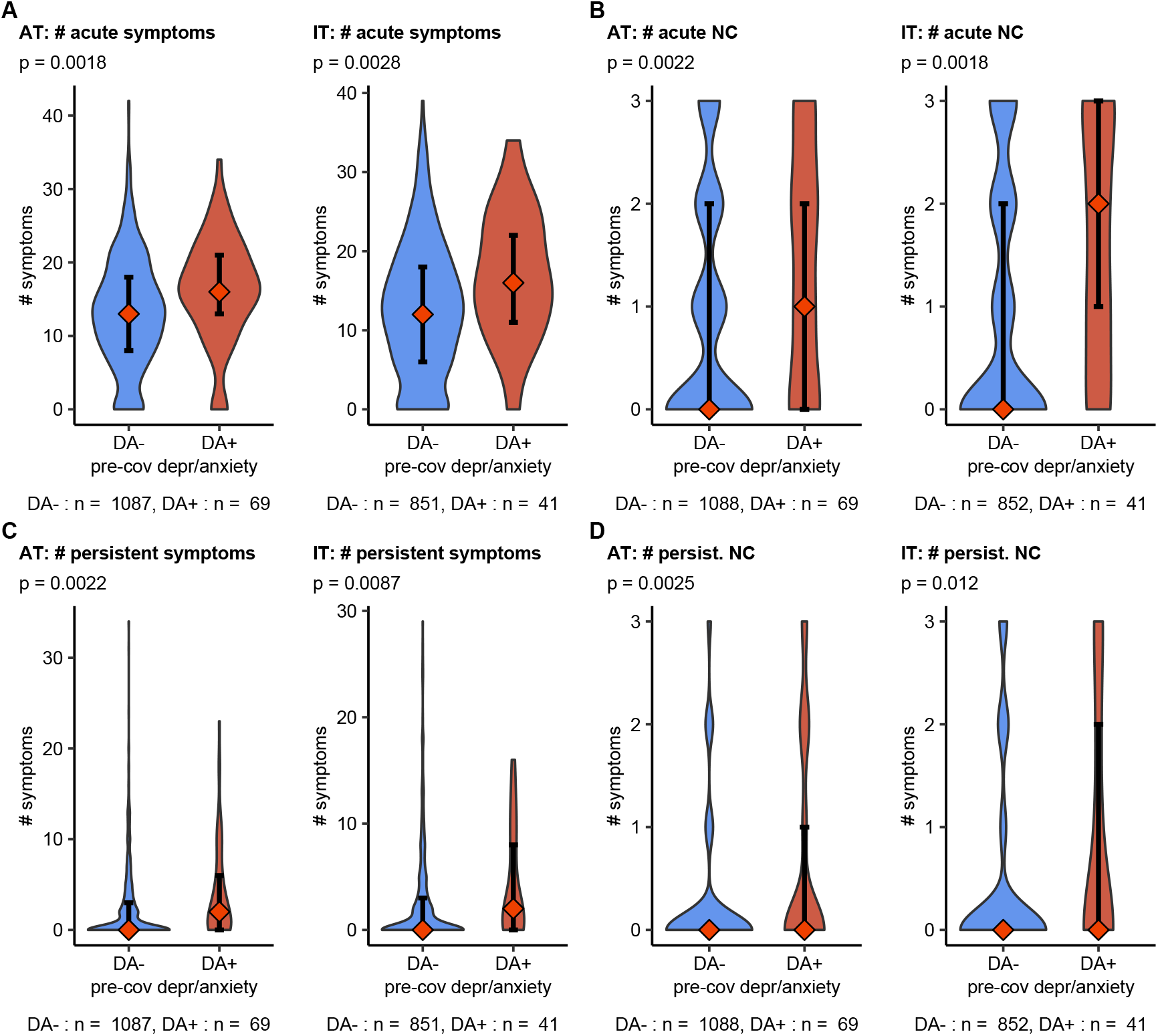
Depression or anxiety before COVID-19 and COVID-19 symptom burden. Association of depression or anxiety before COVID-19 with the overall number of acute and persistent COVID-19 symptoms and neurocognitive (NC) COVID-19 symptoms was assessed by Mann-Whitney U test. Symptom numbers are presented as violin plots, diamonds with whiskers represent medians with IQRs. p values corrected for multiple comparisons with Benjamini-Hochberg method are shown in plot sub-headings. N numbers of observations are indicated below the plots.

## Discussion

In our binational survey, approximately 20% of non-hospitalized COVID-19 convalescents reported poor overall mental health, reduced quality of life, clinical signs of depression or anxiety at about 3 months post infection. High levels of psychosocial stress, poly-symptomatic acute and post-acute COVID-19 course and acute neurocognitive manifestations (impaired concentration, confusion, forgetfulness) were identified as strong explanatory factors.

So far, mental health disorders following COVID-19 were investigated primarily in hospitalized patients. Signs of at least one psychiatric sequelae (PTSD, depression, anxiety, insomnia and obsessive compulsive symptomatology) was discerned in 56% of inpatients at one month after discharge(24). Anxiety, depression and sleep difficulties were present in approximately one-quarter of hospitalized COVID-19 individuals at the 5 – 12 month follow-ups (5–7). In large-scale studies encompassing both in- and outpatients, COVID-19 was identified as an important risk factor for anxiety, stress-related and depressive adjustment disorders(9) and mental health conditions were ascertained in nearly one-fifth of COVID-19 convalescents(8). Of note, this figure is comparable with the frequency of PHQ-4 positive anxiety (AT: 12.4%, IT: 19.3%) and depression screening (AT: 17.3, IT: 23.2%) in our study cohorts. The variability of the reported rates of depression or anxiety in COVID-19 convalescents could be explained both by the differences in assessment methods and by the differing regional containment policies reflected by the rising frequencies of mental conditions in the general population(3). This may explain the significantly higher prevalence of post-COVID-19 depression and anxiety in the IT than in the AT study cohort, despite the similar frequency of pre-existing mental disorders.

Our results underscore the negative impact of psychosocial stress and acute neurocognitive symptoms such as concentration and memory deficits on the mental health rating, which likely reflects a net influence of the pandemic management measures and the disease itself. The neurocognitive complaints during acute COVID-19 (5,7,9,10,12,25–27) were found frequently accompanied by lower-respiratory, cardiological and neurological symptoms and sleep disorders (7,10,27). and posed unfavorable correlates long COVID risk(10). Herein, the acute neurocognitive features together with high symptom burden of acute COVID-19, multiple acute pain manifestations and sleep problems defined the IR and HR Mental Disorder Risk Clusters of the participants likely to develop a mental health condition in course of the recovery. Such ‘red flags’ of deteriorating mental health present in the first two weeks of COVID-19 may be exploited for early diagnosis and psychological or psychiatric intervention.

Pre-existing depression or anxiety was reported by roughly 5% of the respondents and was linked to mental health deficits during recovery – a phenomenon known from non-COVID-19 medical conditions(28). Concomitantly, the subset with pre-existing depression/anxiety was found to experience a significantly higher burden of acute symptoms and post-acute sequelae. Conspicuously, psychiatric disorders before COVID-19 were described as an age- and other comorbidity-independent risk factor for SARS-CoV-2 infection (8). This indicates that alike chronic somatic diseases, pre-existing mental health conditions may predispose the patient to more severe and poly-symptomatic COVID-19.

Several mechanisms might mediate the bidirectional associations of COVID-19, depression, anxiety and psychosocial stress(29). Protracted systemic inflammation is an important pathogenetic factor in depressive-anxious disorders during COVID-19 convalescence (7,10,24,30–32). Stress being the key co-variate of poor mental health in the study collectives was proposed to modulate anti-SARS-CoV-2 immunity culminating in more severe COVID-19 (33) and to perpetuate the systemic low-grade inflammation(28,33). Other possible mechanisms include direct viral infection of the central nervous system, neuroinflammation, microvascular thrombosis and neurodegeneration(34). The strong association of acute neurocognitive manifestations with poor mental health scoring in the our study suggests that pathobiological processes triggered likely by the pathogen and anti-SARS-CoV-2 immunity early in the disease course may contribute to the mental health deterioration.

The prime strength of our study is a broad palette of variables analyzed in two independently recruited cohorts differing in socioeconomic structure and national containement measures which allowed for identification and validation of common influencing factors. Furthermore, the study cohorts encompassed outpatients only insufficiently characterized so far. The most important study limitation is a possible participants’ selection bias. The respondents showed good mental health before COVID-19, and it is likely that predominantly individuals with severe or persistent symptoms and high health-awareness completed the survey(10). Furthermore, individual variability of the clinical onset – survey time may have affected some of the investigated parameters even though their impact on the mental health rating was minimal. Notably, the respective observation time variable was included in the multi-parameter models (11).

## Conclusions

This study underlines the importance of mental health in the follow-up care of COVID-19 individuals. Psychosocial stress, poly-symptomatic disease and neurocognitive complaints accompanying the acute disease may be regarded as ‘red flags’ of a post-COVID-19 mental disorder. They may prompt clinicians, including general practitioners, to monitor outpatients with COVID-19 more closely for mental health deterioration and identify those who could benefit from early psychological and psychiatric intervention. Additionally, a pre-existing mental health condition may pose a risk factor of more severe COVID-19.

## Data Availability

The datasets used and/or analysed during the current study are available from the corresponding author on reasonable request. Analysis of the psychosocial features is available as an online dashboard (https://im2-ibk.shinyapps.io/mental_health_dashboard). The complete R analysis pipeline is available at https://github.com/PiotrTymoszuk/mental-health-after-COVID-19.

## Declarations

### Ethics approval and consent to participate

Each participant gave a digitally signed informed consent to participate. The study protocol was approved by the institutional review boards of the Medical University of Innsbruck (AT, approval number: 1257/2020) and of the Autonomous Province of Bolzano – South Tyrol (IT: 0150701).

### Consent for publication

Not applicable.

### Availability of data and materials

The datasets used and/or analysed during the current study are available from the corresponding author on reasonable request. Analysis of the psychosocial features is available as an online dashboard (*https://im2-ibk.shinyapps.io/mental_health_dashboard*). The complete R analysis pipeline is available at *https://github.com/PiotrTymoszuk/mental-health-after-COVID-19*.

### Competing interests

PT owns his data science enterprise, Data Analytics as a Service Tirol. PT has also received an honorarium for the study data management, curation and analysis and minor manuscript work. The author has no other competing interests to declare. Other authors have no competing interests to declare.

### Funding

The study was funded by the Research Fund of the State of Tyrol, Austria (Project GZ 71934, JLR).

### Author’s contribution

KH, MG, GW, RH, HB, SS, VR, AP, AH, GP, CW, BRW, RH, SK, JLR and BSU designed the study. KH, DA, SS, AP, VR, MG, AB, KK, TS, IT, BP, CW, HB, GP collected the data. KH, PT, DA performed data analysis. PT, DA, KH, RH, BSU and JLR interpreted the data. PT, DA, KH, BSU and JLR wrote the manuscript. All authors critically reviewed the final version of the manuscript.

## Acknowledgments

We acknowledge the commitment of the participants, healthcare administration and general practitioners to the study and daily management of the COVID-19 pandemic.

## Notes

### Clinical Trial

NCT04661462

### Author Declarations

Each participant gave a digitally signed informed consent to participate. The study protocol was approved by the institutional review boards of the Medical University of Innsbruck (AT, approval number: 1257/2020) and of the Autonomous Province of Bolzano - South Tyrol (IT: 0150701).

## References

1. Martin A, Rief W, Klaiberg A, Braehler E. Validity of the Brief Patient Health Questionnaire Mood Scale (PHQ-9) in the general population. Gen Hosp Psychiatry [Internet]. 2006 Jan [cited 2021 Jun 24];28(1):71–7. Available from: https://pubmed.ncbi.nlm.nih.gov/16377369/

2. Löwe B, Decker O, Müller S, Brähler E, Schellberg D, Herzog W, et al. Validation and standardization of the generalized anxiety disorder screener (GAD-7) in the general population. Med Care [Internet]. 2008 Feb [cited 2021 Jun 24];46(3):266–74. Available from: https://pubmed.ncbi.nlm.nih.gov/18388841/

3. Pieh C, Budimir S, Probst T. The effect of age, gender, income, work, and physical activity on mental health during coronavirus disease (COVID-19) lockdown in Austria. J Psychosom Res. 2020 Sep 1;136:110186.

4. Nasserie T, Hittle M, Goodman SN. Assessment of the Frequency and Variety of Persistent Symptoms Among Patients With COVID-19: A Systematic Review. JAMA Netw open [Internet]. 2021 May 3 [cited 2021 Jun 24];4(5):e2111417. Available from: http://www.ncbi.nlm.nih.gov/pubmed/34037731

5. Huang C, Huang L, Wang Y, Li X, Ren L, Gu X, et al. 6-month consequences of COVID-19 in patients discharged from hospital: a cohort study. Lancet [Internet]. 2021 Jan 16 [cited 2021 May 20];397(10270):220–32. Available from: https://covid19.who.int/

6. Huang L, Yao Q, Gu X, Wang Q, Ren L, Wang Y, et al. 1-year outcomes in hospital survivors with COVID-19: a longitudinal cohort study. Lancet [Internet]. 2021 Aug 28 [cited 2021 Sep 13];398(10302):747–58. Available from: https://linkinghub.elsevier.com/retrieve/pii/S0140673621017554

7. Evans RA, McAuley H, Harrison EM, Shikotra A, Singapuri A, Sereno M, et al. Physical, cognitive and mental health impacts of COVID-19 following hospitalisation – a multi-centre prospective cohort study. medRxiv [Internet]. 2021 Mar 25 [cited 2021 Sep 20];2021.03.22.21254057. Available from: https://doi.org/10.1101/2021.03.22.21254057

8. Taquet M, Luciano S, Geddes JR, Harrison PJ. Bidirectional associations between COVID-19 and psychiatric disorder: retrospective cohort studies of 62 354 COVID-19 cases in the USA. The Lancet Psychiatry [Internet]. 2021 Feb 1 [cited 2021 Jun 24];8(2):130–40. Available from: https://pubmed.ncbi.nlm.nih.gov/33181098/

9. Al-Aly Z, Xie Y, Bowe B. High-dimensional characterization of post-acute sequalae of COVID-19. Nature [Internet]. 2021 Apr 22 [cited 2021 May 24];1–6. Available from: http://www.ncbi.nlm.nih.gov/pubmed/33887749

10. Sahanic S, Tymoszuk P, Ausserhofer D, Rass V, Pizzini A, Nordmeyer G, et al. Phenotyping of acute and persistent COVID-19 features in the outpatient setting: exploratory analysis of an international cross-sectional online survey. medRxiv [Internet]. 2021 Aug 7 [cited 2021 Aug 9];2021.08.05.21261677. Available from: https://doi.org/10.1101/2021.08.05.21261677

11. Health after COVID-19 in Tyrol. Mental Health after COVID-19 in Tyrol [Internet]. [cited 2021 Sep 9]. Available from: https://im2-ibk.shinyapps.io/mental_health_dashboard/

12. Sudre CH, Murray B, Varsavsky T, Graham MS, Penfold RS, Bowyer RC, et al. Attributes and predictors of long COVID. Nat Med [Internet]. 2021 [cited 2021 Apr 25];27(4). Available from: https://pubmed.ncbi.nlm.nih.gov/33692530/

13. Löwe B, Wahl I, Rose M, Spitzer C, Glaesmer H, Wingenfeld K, et al. A 4-item measure of depression and anxiety: Validation and standardization of the Patient Health Questionnaire-4 (PHQ-4) in the general population. J Affect Disord [Internet]. 2010 Apr [cited 2021 Jun 24];122(1–2):86–95. Available from: https://pubmed.ncbi.nlm.nih.gov/19616305/

14. Gräfe K, Zipfel S, Herzog W, Löwe B. Screening psychischer störungen mit dem “Gesundheitsfragebogen für Patienten (PHQ-D)”. Ergebnisse der Deutschen validierungsstudie. Diagnostica [Internet]. 2004 Oct 1 [cited 2021 Sep 10];50(4):171–81. Available from: https://econtent.hogrefe.com/doi/abs/10.1026/0012-1924.50.4.171

15. Wickham H, Averick M, Bryan J, Chang W, McGowan L, François R, et al. Welcome to the Tidyverse. J Open Source Softw. 2019 Nov 21;4(43):1686.

16. Wickham H. ggplot2: Elegant Graphics for Data Analysis [Internet]. 1st ed. New York: Springer-Verlag; 2016. Available from: https://ggplot2.tidyverse.org

17. Benjamini Y, Hochberg Y. Controlling the False Discovery Rate: A Practical and Powerful Approach to Multiple Testing. J R Stat Soc Ser B. 1995 Jan 1;57(1):289–300.

18. Breiman L. Random forests. Mach Learn [Internet]. 2001 Oct [cited 2021 Feb 12];45(1):5–32. Available from: https://link.springer.com/article/10.1023/A:1010933404324

19. Kuhn M. Building predictive models in R using the caret package. J Stat Softw. 2008;28(5):1–26.

20. Croux C, Filzmoser P, Oliveira MR. Algorithms for Projection-Pursuit robust principal component analysis. Chemom Intell Lab Syst. 2007 Jun 15;87(2):218–25.

21. Kohonen T. Self-Organizing Maps [Internet]. Berlin, Heidelberg: Springer Berlin Heidelberg; 1995 [cited 2021 Sep 2]. (Springer Series in Information Sciences; vol. 30). Available from: http://link.springer.com/10.1007/978-3-642-97610-0

22. Vesanto J, Alhoniemi E. Clustering of the self-organizing map. IEEE Trans Neural Networks. 2000 May;11(3):586–600.

23. Borenstein M, Hedges L V., Higgins JPT, Rothstein HR. A basic introduction to fixed-effect and random-effects models for meta-analysis. Res Synth Methods [Internet]. 2010 Apr [cited 2021 Jul 30];1(2):97–111. Available from: https://pubmed.ncbi.nlm.nih.gov/26061376/

24. Mazza MG, De Lorenzo R, Conte C, Poletti S, Vai B, Bollettini I, et al. Anxiety and depression in COVID-19 survivors: Role of inflammatory and clinical predictors. Brain Behav Immun [Internet]. 2020 Oct 1 [cited 2021 Sep 10];89:594–600. Available from: https://pubmed.ncbi.nlm.nih.gov/32738287/

25. Blomberg B, Mohn KGI, Brokstad KA, Zhou F, Linchausen DW, Hansen BA, et al. Long COVID in a prospective cohort of home-isolated patients. Nat Med [Internet]. 2021 [cited 2021 Aug 5]; Available from: https://pubmed.ncbi.nlm.nih.gov/34163090/

26. Goërtz YMJ, Van Herck M, Delbressine JM, Vaes AW, Meys R, Machado FVC, et al. Persistent symptoms 3 months after a SARS-CoV-2 infection: the post-COVID-19 syndrome? ERJ Open Res [Internet]. 2020 Oct [cited 2021 Aug 5];6(4):00542–2020. Available from: https://pubmed.ncbi.nlm.nih.gov/33257910/

27. Davis HE, Assaf GS, McCorkell L, Wei H, Low RJ, Re’em Y, et al. Characterizing long COVID in an international cohort: 7 months of symptoms and their impact. EClinicalMedicine [Internet]. 2021 Jul [cited 2021 Aug 5];101019. Available from: https://pubmed.ncbi.nlm.nih.gov/34308300/

28. Best C, Eckhardt-Henn A, Tschan R, Dieterich M. Psychiatric morbidity and comorbidity in different vestibular vertigo syndromes: Results of a prospective longitudinal study over one year. J Neurol [Internet]. 2009 Jan [cited 2021 Sep 10];256(1):58–65. Available from: https://pubmed.ncbi.nlm.nih.gov/19221849/

29. Postolache TT, Benros ME, Brenner LA. Targetable Biological Mechanisms Implicated in Emergent Psychiatric Conditions Associated with SARS-CoV-2 Infection [Internet]. Vol. 78, JAMA Psychiatry. American Medical Association; 2021 [cited 2021 Sep 10]. p. 353–4. Available from: https://pubmed.ncbi.nlm.nih.gov/32735332/

30. Hu Y, Chen Y, Zheng Y, You C, Tan J, Hu L, et al. Factors related to mental health of inpatients with COVID-19 in Wuhan, China. Brain Behav Immun [Internet]. 2020 Oct 1 [cited 2021 Sep 10];89:587–93. Available from: https://pubmed.ncbi.nlm.nih.gov/32681866/

31. Parker C, Shalev D, Hsu I, Shenoy A, Cheung S, Nash S, et al. Depression, Anxiety, and Acute Stress Disorder Among Patients Hospitalized With Coronavirus Disease 2019: A Prospective Cohort Study. Psychosomatics [Internet]. 2020 [cited 2021 Sep 10];62(2). Available from: https://pubmed.ncbi.nlm.nih.gov/33198962/

32. Mazza MG, Palladini M, De Lorenzo R, Magnaghi C, Poletti S, Furlan R, et al. Persistent psychopathology and neurocognitive impairment in COVID-19 survivors: Effect of inflammatory biomarkers at three-month follow-up. Brain Behav Immun [Internet]. 2021 May 1 [cited 2021 Sep 10];94:138–47. Available from: https://pubmed.ncbi.nlm.nih.gov/33639239/

33. Peters EMJ, Schedlowski M, Watzl C, Gimsa U. Can Stress Interact with SARS-CoV-2? A Narrative Review with a Focus on Stress-Reducing Interventions that may Improve Defence against COVID-19 [Internet]. Vol. 71, PPmP Psychotherapie Psychosomatik Medizinische Psychologie. Georg Thieme Verlag; 2021 [cited 2021 Sep 10]. p. 61–71. Available from: https://pubmed.ncbi.nlm.nih.gov/33440452/

34. Nalbandian A, Sehgal K, Gupta A, Madhavan M V., McGroder C, Stevens JS, et al. Postacute COVID-19 syndrome [Internet]. Vol. 27, Nature Medicine. Nature Research; 2021 [cited 2021 Jul 1]. p. 601–15. Available from: https://doi.org/10.1038/s41591-021-01283-z

