## Supplementary Material for "Who is at risk of poor mental health following COVID-19 outpatient management?"

Health after COVID-19 in Tyrol study team

2021-09-22

**Data availability** **4**

**Supplementary Tables** **5**

**Supplementary Figures** **11**

**References** **28**

### Supplementary Methods

#### Study design and participants

The multi-center binational online survey study 'Health after COVID-19 in Tyrol' (ClinicalTrials.gov: NCT04661462) was conducted between the 30$^{th}$ September 2020 and 11$^{th}$ July 20221 in two independently recruited cohorts in Tyrol/Austria (AT) and South Tyrol/Italy (IT).[1] The study cohorts encompassed residents of the study regions aged $\geq$ 16 (AT) or $\geq$ 18 years (IT) who experienced a laboratory-confirmed SARS-CoV2 infection (PCR or seropositivity). The respondents with a minimum observation time of <28 days between the infection diagnosis and survey completion or hospitalized because of COVID-19 were excluded from the analysis. The scheme of study and analysis enrollment is depicted in **Figure 1**. The participants were invited by a public media call (both cohorts) or by their general practitioners (IT).

The study was conducted in accordance with the Declaration of Helsinki as well as the national and European data policies. Each participant gave a digitally signed informed consent to participate. The study protocol was approved by the institutional review boards of the Medical University of Innsbruck (AT, approval number: 1257/2020) and of the Autonomous Province of Bolzano - South Tyrol province (IT, 0150701).

#### Measures, definitions and data transformation

The detailed description of the questionnaire is provided by Sahanic et al.[1] In brief, dates of the study completion and SARS-CoV2 infection diagnosis, data on biometry (weight, height), demographics (age, sex), pre-existing co-morbidities, socioeconomic status (residence region, mother tongue, employment status and profession), smoking history and COVID-19 relevant medication, symptom duration (44 items), symptomatic therapy and course of SARS-CoV2 infection, recovery duration and status as well as mental health and psychosocial stress following the disease were queried. The complete list of features analyzed in the current report is presented in **Supplementary Table S1** and the baseline characteristic of the study collectives is shown in **Table 1** and **Supplementary Table S2**.

Observation time defined as a time interval between the diagnosis of SASR-CoV-2 infection (positive test) and the survey completion was stratified as follows: up to 60 days, 61 - 120 days, 121 – 180 days, more than 180 days. Self-reported COVID-19 symptoms were retrospectively assigned to the following duration classes: absent, present for 1 – 3 days, $\leq$ 1 week, $\leq$ 2 weeks, $\leq$ 4 weeks, $\leq$ 3 months, $\leq$ 6 months and > 6 months. Symptoms were classified as acute complaints present during the first 2 weeks after clinical onset and persistent symptoms present for $\geq$ 4 weeks.[1] Confusion, impaired concentration and forgetfulness were classified as 'neurocognitive symptoms'. For modeling, the overall number of overall acute symptoms was stratified by quartiles (Q1 - Q4) and the overall persistent symptom count by median and 75$^{th}$ percentile . The number of acute neuro-cognitive symptoms was stratified by median and 75$^{th}$ percentile and for persistent neuro-cognitive manifestations coded as an index present/absent variable (**Supplementary Table S1** and **Table 2**).

Depression/anxiety before SARS-CoV2 infection, pre-existing sleep disorders, acute COVID-19 perception (common cold-, influenza-, gastroenteritis-like or unique/not experienced before), symptom relapse, complete convalescence, rehabilitation need and percent physical performance loss following COVID-19 were surveyed as single question items each. For modeling, the physical performance loss was stratified as 0 - 25%, 26 - 50%, 51% - 75% and 76 - 100%.

Self-perceived overall mental health (OMH) and quality of life (QoL) were assessed as single questions ('excellent', 'good', 'fair', 'poor', scored: 0, 1, 2, 3). Anxiety/depression following COVID-19 at time of study completion were investigated using PHQ-4 module (two questions each, possible answers: 'never', 'some days', 'over 50% of days', 'almost every day', scoring: 0, 1, 2, 3 points). Clinical signs of depression (DPR) or anxiety (ANX) were defined with the cutoffs of $\geq$ 3 point sum.[2] Psychosocial stress was measured with a modified 7 item (answers: 'no', 'little', 'some', 'a lot', scored: 0, 1, 2, 3) PHQ stress module,[3–6] without items on weight, sexuality and past traumatic/serious events; the item on worries/dreams was adapted to

COVID-19. The stress scoring was re-coded as quartile strata encompassing 0 – 2, 3 – 4, 5 – 6, 7 – 21 points. Substantial psychosocial stress was defined by a $\geq 7$ point cutoff.

#### Statistical analysis

##### Data transformation, visualization, descriptive statistic and hypothesis testing

The study variables were transformed, analyzed and visualized with R version 4.0.5 with *tidyverse*,[7,8] *cowplot*[9] and *ggvenn* packages.

For categorical variables, numbers and percents of complete answers are presented. As most of the analyzed numeric features had a discrete or non-normal distribution as checked by Shapiro-Wilk test, medians, interquartile ranges (IQR) and feature ranges are presented. To compare differences in distribution of categorical features, $\chi^2$ test was applied. To assess significance of differences in numeric variables between groups, U or Kruskal-Wallis test was used, as appropriate. Co-occurrence of two categorical variables was expressed as Cohen's kappa statistics (function *Kappa()*, package *vcd*), whose significance was assessed by Z test. P values were corrected for multiple comparisons with Benjamini-Hochberg method.[10] The set of tools used for descriptive statistics and hypothesis testing is available from https://github.com/PiotrTymoszuk/counting-tools.

##### Random forest modeling of mental health and quality of life scoring

Multi-parameter random forest regression models[11] describing the scoring of OMH, QoL, ANX and DPR separately in the Austria/Tyrol and Italy/South Tyrol cohorts as functions of 145 independent parameters (**Supplementary Table S1**) were constructed and verified in by 10-fold cross-validation using *caret* package (function *train()*, *mtry* argument specifying the number of random tree models was set to 500).[12] Of note, to account for possible recall bias of acute COVID-19 symptoms and effects of convalescence time on the scoring and frequency of clinical signs of mental health disorders, the stratified observation time variable was included in the random forest modeling procedure. The model fits to the training data set are presented in **Supplementary Figures S5 - S4** and were assessed by Spearman regression and mean absolute error (MAE) values for the training and cross-validation data sets.

To discern the features with the largest effect on the OMH, QoL, ANX and DPR scoring each, differences in mean squared error ($\Delta$MSE) associated with the model components were extracted using *importance()* function on the final models developed by *caret* (**Supplementary Figures S5 - S4**).[11,13] To identify common factors with the greatest impact on the combined mental health and quality of life scoring, the normalized values of $\Delta$MSE of the OMH, QoL, ANX and DPR scoring for each model component were subjected to centered principal component analysis (PCA) using *PCAproj()* function from *pcaPP* package.[14] The features with the 10 largest PCA loadings were further for univariable modeling and clustering .

##### Univariable modeling

Correlation of the OMH, QoL, ANX and DPR scoring and the most influential factors identified by the random forest technique was assessed by age- and sex-weighted Poisson regression (generalized linear modeling, log link function). The frequency weights for the Austria/Tyrol and Italy/South Tyrol cohort were based on the age and sex distribution of COVID cases in Tyrol[15] and Italy,[16] respectively[1]. Significance of model estimates and their 95% confidence intervals were determined with Wald Z test. P values were corrected for multiple comparisons by Benjamini-Hochberg method.[10] The correlation was deemed significant when significant association was present in both study cohorts. Model estimate extraction and visual quality control was accomplished with home-developed R tools (https://github.com/PiotrTymoszuk/lm_qc_tools). For complete univariable modeling results, see: **Supplementary Table S3**.

Pooled Austria/Italy $\beta$ estimates referenced to in the text and presented in **Supplementary Table S4** were calculated using inversed variance method and *meta* package.[17,18]

##### Definition of the mental disorder risk clusters

The study participants, separately in the Austian and Italian cohort, were clustered using a two-step combined self-organizing map (SOM) and hierarchical clustering algorithm[19,20] in respect to the most influential factors affecting the combined mental health and quality of life scoring identified by multi-parameter random forest modeling and PCA. In the first step, participants were assigned to the nodes of 11 × 11 unit hexagonal grid with the Jaccard distance measure between the participants. The grid size was estimated with the $5 \times \sqrt{N}$ formula, where N is the number of observations in the smaller Italian data set, as proposed by Vesanto et al.[21] SOM assignment was accomplished with the tools provided by *kohonen* package and home-developed wrappers (https://github.com/PiotrTymoszuk/SOM_tools). The SOM training process is visualized in **Figure S8A**. In the second step, SOM nodes were subjected to hierarchical clustering with Ward D2 method and Euclidean distance measure. The optimal cluster number (k = 3) was determined by the bend of the within sum-of-squares and visual analysis of the dendrograms (**Supplementary Figure S8BC**). The hierarchical clustering was done with the base *hclust()* function and home-developed wrappers for clustering quality control and visualization (https://github.com/PiotrTymoszuk/cluster_tools).

### Data availability

As this study is still ongoing, the complete data will be made available on a serious request to the corresponding author and made publicly available after the completion. Analysis of the psychosocial features is available as an online *R shiny* dashboard at Mental Health after COVID-19 in Tyrol (https://im2-ibk.shinyapps.io/mental_health_dashboard/).[6] The R analysis pipeline is available at https://github.com/PiotrTymoszuk/mental-health-after-COVID-19.

#### Supplementary Tables

Table S1: **Survey variables used for construction of random forest models.**. The table is available online.

Table S2: **Supplementary characteristic of the study cohorts.**
AT: Austria/Tyrol cohort, IT: Italy/South Tyrol cohort, Test: statistical test used for the AT vs IT comparison, Significance: test p value corrected for multiple comparisons with Benjamini-Hochberg method.

| Variable | AT | IT | Test | Significance |
|---|---|---|---|---|
| Time between survey and diagnosis | median(IQR) = 79 (40 - 175)<br>range = 28 - 400<br>n = 1157 | median(IQR) = 96 (60 - 138)<br>range = 28 - 387<br>n = 893 | U | p = 1.2e-07 |
| | up to 60 days: 42.5% (492)<br>61 - 120 days: 20.2% (234)<br>121 - 180 days: 14.2% (164)<br>more than 180 days: 23.1% (267)<br>n = 1157 | up to 60 days: 25.8% (230)<br>61 - 120 days: 35.4% (316)<br>121 - 180 days: 21.6% (193)<br>more than 180 days: 17.2% (154)<br>n = 893 | $\chi^2$ | p = 1.3e-22 |
| Survey completion | fall 2020: 63.4% (734)<br>winter/spring 2021: 36.6% (423)<br>n = 1157 | fall 2020: 4.37% (39)<br>winter/spring 2021: 95.6% (854)<br>n = 893 | $\chi^2$ | p = 4.2e-163 |
| Region | capital: 19.9% (230)<br>non-capital: 80.1% (927)<br>n = 1157 | capital: 56.6% (505)<br>non-capital: 43.4% (388)<br>n = 893 | $\chi^2$ | p = 5.6e-65 |
| Native language | German: 100% (1157)<br>Italian: 0% (0)<br>Ladin: 0% (0)<br>Other: 0% (0)<br>n = 1157 | German: 55.3% (493)<br>Italian: 36.7% (327)<br>Ladin: 6.5% (58)<br>Other: 1.57% (14)<br>n = 892 | $\chi^2$ | p = 4.5e-138 |
| Employment sector | other: 18.7% (214)<br>gastronomy/tourism: 8.82% (101)<br>health services: 25.9% (296)<br>food trade: 2.18% (25)<br>public transportation: 0.786% (9)<br>emergency services: 2.1% (24)<br>construction: 2.97% (34)<br>administration/office: 19.4% (222)<br>industry: 5.68% (65)<br>agriculture: 0.961% (11)<br>education: 12.6% (144)<br>n = 1145 | other: 18.1% (157)<br>gastronomy/tourism: 8.29% (72)<br>health services: 20.1% (175)<br>food trade: 1.84% (16)<br>public transportation: 0.575% (5)<br>emergency services: 0% (0)<br>construction: 3.11% (27)<br>administration/office: 28.2% (245)<br>industry: 4.95% (43)<br>agriculture: 1.5% (13)<br>education: 13.3% (116)<br>n = 869 | $\chi^2$ | p = 7.3e-06 |
| Diabetes | 1.56% (18)<br>n = 1157 | 0.784% (7)<br>n = 893 | $\chi^2$ | ns |

Table S2: **Supplementary characteristic of the study cohorts.**
AT: Austria/Tyrol cohort, IT: Italy/South Tyrol cohort, Test: statistical test used for the AT vs IT comparison, Significance: test p value corrected for multiple comparisons with Benjamini-Hochberg method. *(continued)*

| Variable | AT | IT | Test | Significance |
|---|---|---|---|---|
| Gastrointestinal disease | 1.56% (18)<br>n = 1157 | 1.01% (9)<br>n = 893 | $\chi^2$ | ns |
| Malignancy | 2.42% (28)<br>n = 1157 | 2.91% (26)<br>n = 893 | $\chi^2$ | ns |
| > 2 respiratory infections per year | 4.41% (51)<br>n = 1157 | 2.91% (26)<br>n = 893 | $\chi^2$ | ns |
| > 2 bacterial infections per year | 3.89% (45)<br>n = 1157 | 1.34% (12)<br>n = 893 | $\chi^2$ | p = 0.0017 |
| Hair loss | 13.7% (158)<br>n = 1157 | 14.8% (132)<br>n = 893 | $\chi^2$ | ns |
| Weight loss | none: 52.5% (604)<br>mild: 16.3% (188)<br>moderate: 26% (299)<br>severe: 5.21% (60)<br>n = 1151 | none: 60.4% (538)<br>mild: 15.6% (139)<br>moderate: 19.6% (174)<br>severe: 4.38% (39)<br>n = 890 | $\chi^2$ | p = 0.0025 |
| Complete convalescence | 54% (624)<br>n = 1155 | 63.3% (563)<br>n = 889 | $\chi^2$ | p = 6.6e-05 |
| Physical performance loss | 0 - 25%: 74.7% (860)<br>26 - 50%: 17.5% (202)<br>51% - 75%: 6.26% (72)<br>76 - 100%: 1.48% (17)<br>n = 1151 | 0 - 25%: 76.2% (674)<br>26 - 50%: 16.4% (145)<br>51% - 75%: 6.11% (54)<br>76 - 100%: 1.24% (11)<br>n = 884 | $\chi^2$ | ns |
| Subjective need for rehabilitation | 17% (196)<br>n = 1153 | 13.2% (117)<br>n = 888 | $\chi^2$ | p = 0.033 |

Table S3: **Results of univariate modeling for the most influential mental health scoring factors.** Top 10 factors with the largest impact on the net mental health scoring were determined by random forest prediction and principal component analysis as presented in Figure 2. Their correlation with overall mental health (OMH), quality of life (QoL), anxiety (ANX) and depression (DPR) scoring was investigated by univariate sex- and age-weighted Poisson regression. P values were corrected for multiple testing with Benjamini-Hochberg method. The table is available online.

Table S4: **Pooled Austria/Italy cohort results of univariate modeling for the most influential mental health scoring factors.**
Pooled Austria/Italy $\beta$ estimates for the factors with the largest impact on the net mental health scroring were calculated with inverse variance method. P values were corrected for multiple testing with Benjamini-Hochberg method. The table is available online.

Table S5: **Differences in frequency of the survey items not used for cluster definition between the mental disorder risk clusters.**
Cluster: LR - low risk, IR - intermediate risk, HR - high risk cluster, N: number of observations assigned to the cluster, Significance: p values obtained by $\chi^2$ test corrected for multiple testing with Benjamini-hochber method. The table is available online.

#### Supplementary Figures

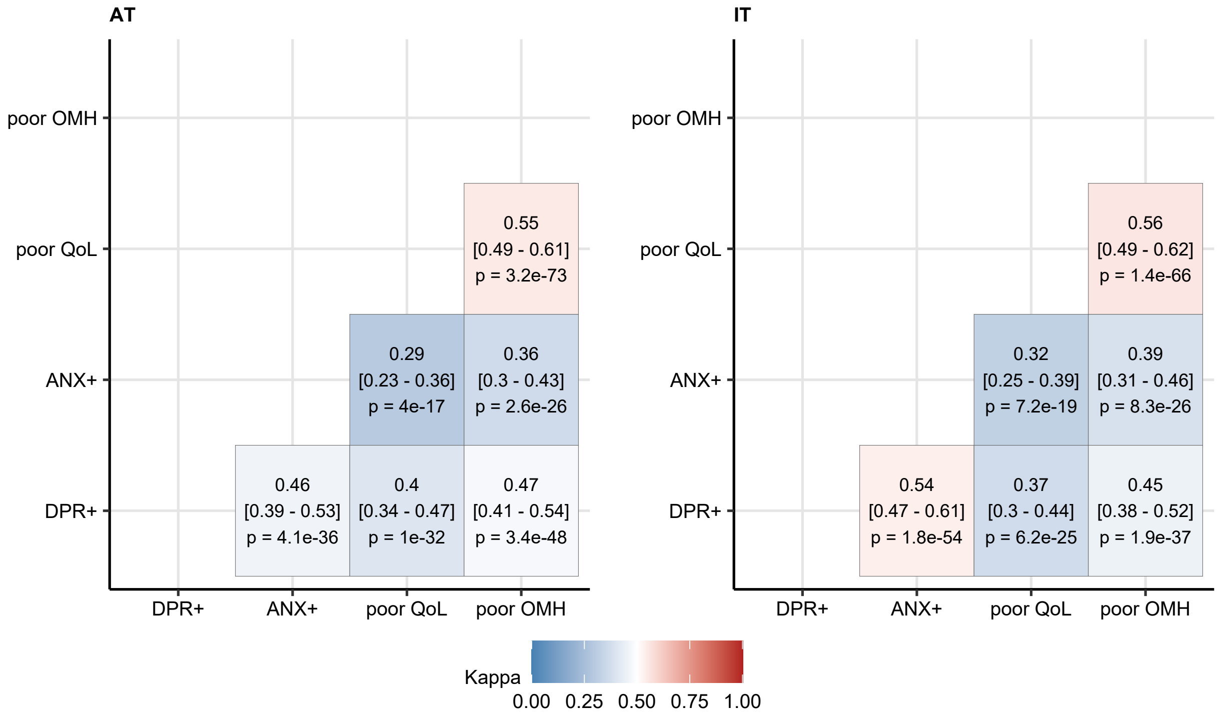

Figure S1: Co-occurrence of depression, anxiety, low rating of overall mental health and quality of life following COVID-19..

**Supplementary Figure S1. Co-occurrence of depression, anxiety, low rating of overall mental health and quality of life following COVID-19.**

Co-occurrence of positive depression screening (DPR+), positive anxiety screening (ANX+), low rating of self-perceived overall mental health (OMH, rated as 'fair' or 'poor') and low rating of self-perceived quality of life (QoL, rated as 'fair' or 'poor') was measured with Cohen's kappa statistic and presented as a heat map. Statistical significance was determined by Z test and p values were corrected for multiple comparisons with Benjamini-Hochberg method. Tile color corresponds with the kappa value. Kappa values with 95% confidence intervals and p values are presented in the plot.

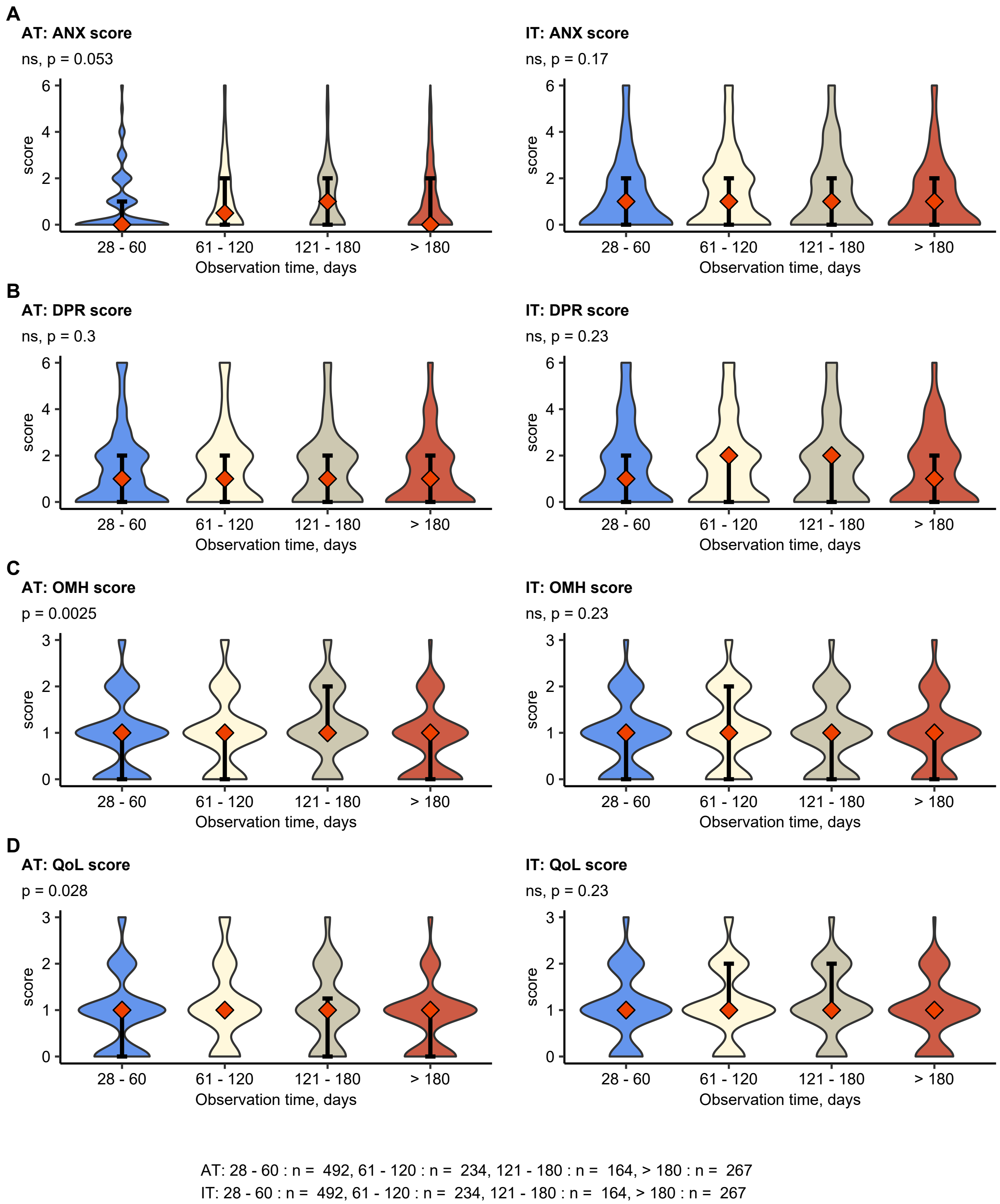

Figure S2: Diagnosis-to-survey time and and mental health scoring.

**Supplementary Figure S2. Diagnosis-to-survey time and and mental health scoring.**
Association of the observation time (SARS-CoV2-2 infection diagnosis to survey completion) with anxiety

(ANX) (**A**), depression (DPR) (**B**), overall mental health (OMH) (**C**) and quality of life (QoL) (**D**) scoring assessed by Kruskal-Wallis test. The scoring is presented as violin plots, diamonds with whiskers represent medians with IQRs. P values corrected for multiple comparisons with Benjamini-Hochberg method are shown in plot sub-headings. N numbers of observations are indicated below the plot.

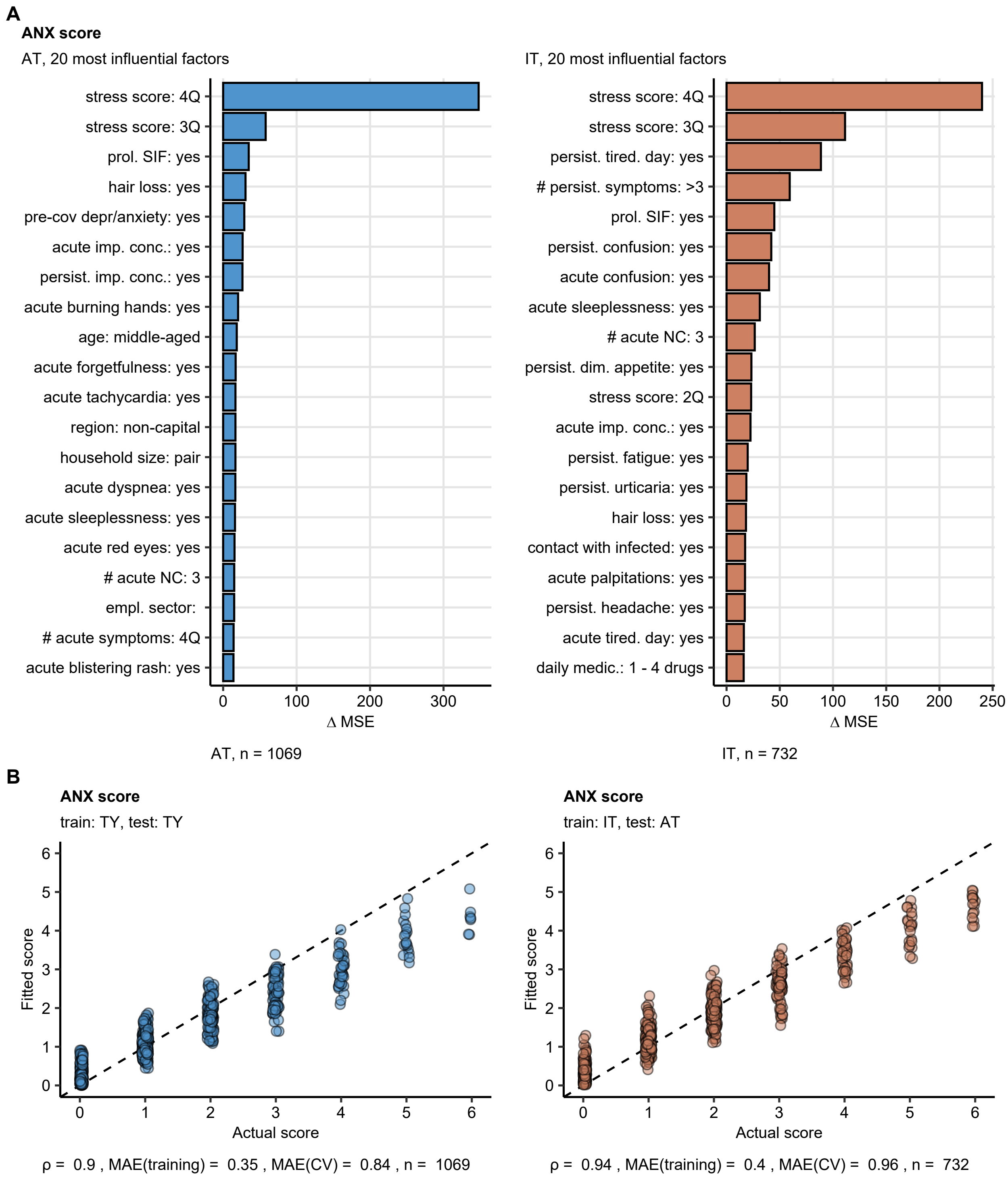

Figure S3: Construction and performance of the anxiety scoring random forest models.

**Supplementary Figure S3. Construction and performance of the anxiety scoring random forest models.**

Random forest models fitting 145 survey variables (**Supplementary Table S1**) to the anxiety scoring in the Austria/Tyrol (AT) and Italy/South Tyrol (IT) cohorts were constructed and validated by the 10-fold cross-validation (CV) technique.

(**A**) Top 20 most influential factors contributing to the improvement of to model fit measured as difference in mean squared error ($\Delta$MSE). N numbers of observations are indicated below the plot.

(**B**) Fitted versus true scoring values in the Austria/Tyrol and Italy/South Tyrol cohorts. Spearman's $\rho$ correlation coefficients, means absolute errors (MAE) for the whole-cohort and cross-validation data sets and n numbers of observations are indicated below the plot.

imp.: impaired, pre-cov depr/anxiety: depression or anxiety before COVID-19, tired. day: tiredness at day, prol.: prolonged, SIF: severe illness feeling, #: number, NC: neurocognitive symptoms, GP: general practitioner, persist.: persistent, dim.: diminished, 2Q, 3Q, 4Q: $2^{nd}$, $3^{rd}$, $4^{th}$ quartile, empl.: employment, conc.: concentration, daily medic.: daily medication, # cov in household: number of COVID-19 cases in the household, subj. cov percept.: subjective perception of acute COVID-19, pre-cov sleep disord.: sleep disorder before COVID-19.

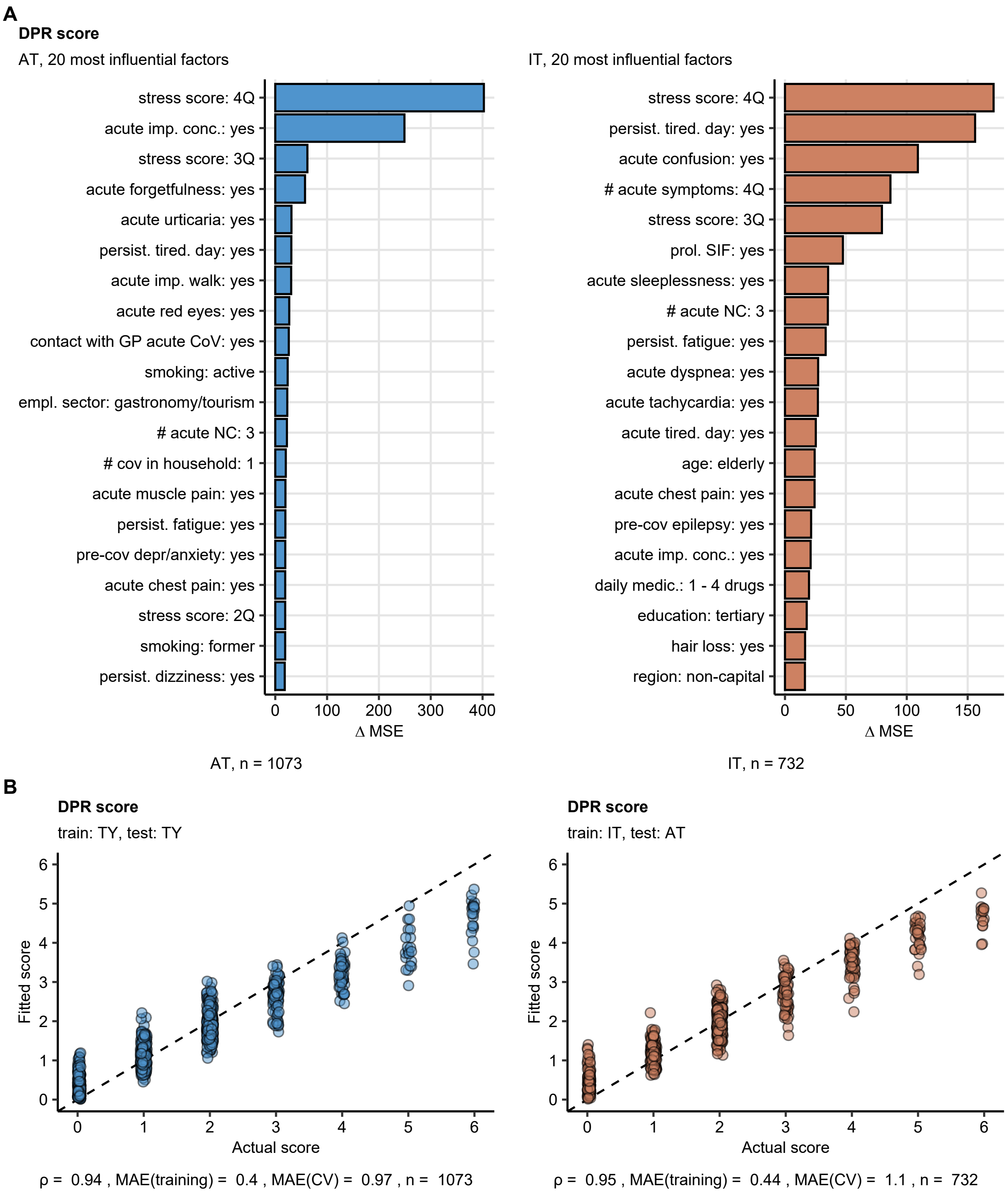

Figure S4: Construction and performance of the depression scoring random forest models.

**Supplementary Figure S4. Construction and performance of the depression scoring random forest models.**

Random forest models fitting 145 survey variables (**Supplementary Table S1**) to the depression scoring in the Austria/Tyrol (AT) and Italy/South Tyrol (IT) cohorts were constructed and validated by the 10-fold cross-validation (CV) technique.

(**A**) Top 20 most influential factors contributing to the improvement of to model fit measured as difference in mean squared error (ΔMSE). N numbers of observations are indicated below the plot.

(**B**) Fitted versus true scoring values in the Austria/Tyrol and Italy/South Tyrol cohorts. Spearman's $\rho$ correlation coefficients, means absolute errors (MAE) for the whole-cohort and cross-validation data sets and n numbers of observations are indicated below the plot.

imp.: impaired, pre-cov depr/anxiety: depression or anxiety before COVID-19, tired. day: tiredness at day, prol.: prolonged, SIF: severe illness feeling, #: number, NC: neurocognitive symptoms, GP: general practitioner, persist.: persistent, dim.: diminished, 2Q, 3Q, 4Q: $2^{nd}$, $3^{rd}$, $4^{th}$ quartile, empl.: employment, conc.: concentration, daily medic.: daily medication, # cov in household: number of COVID-19 cases in the household, subj. cov percept.: subjective perception of acute COVID-19, pre-cov sleep disord.: sleep disorder before COVID-19.

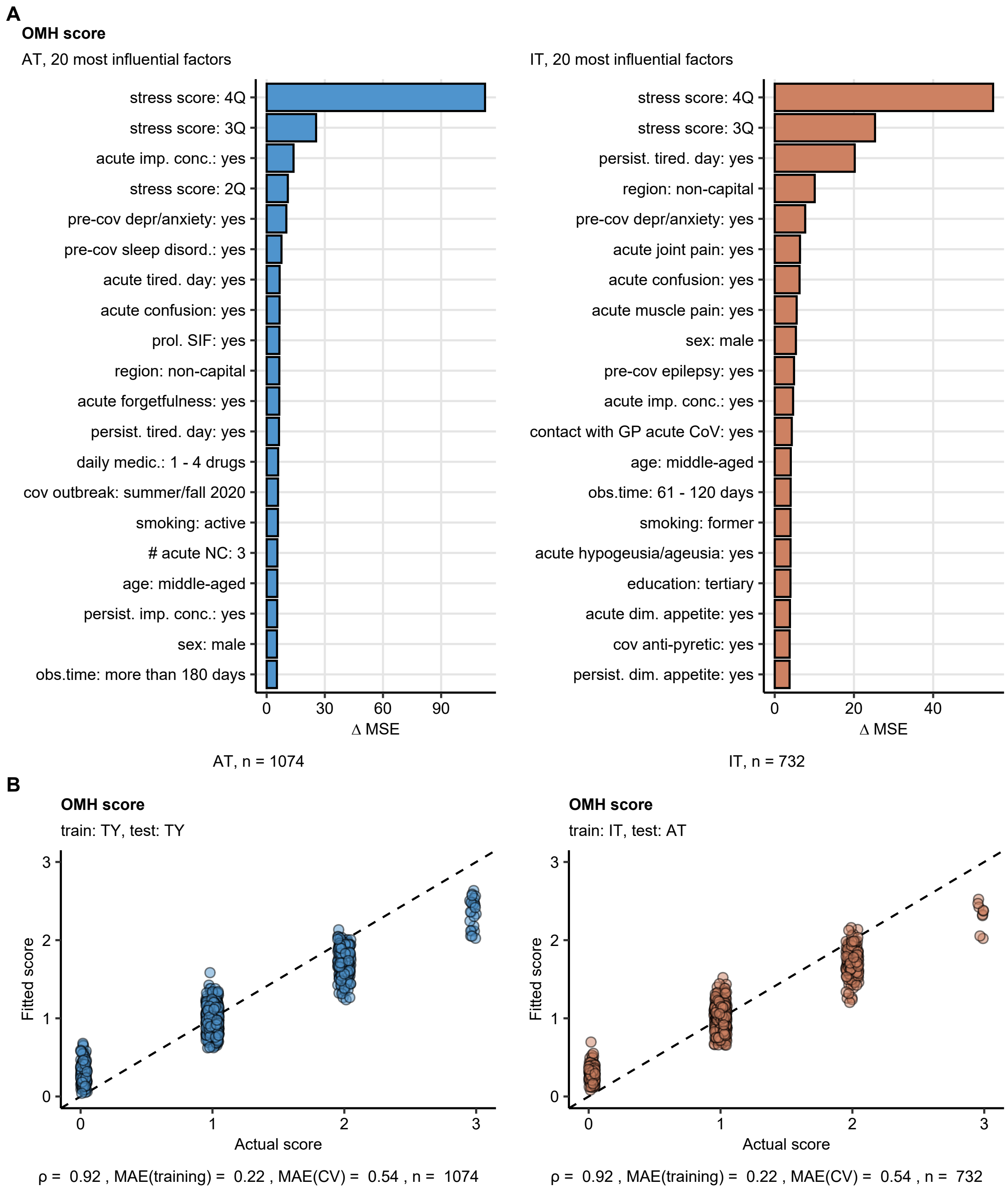

Figure S5: Construction and performance of the overall mental health scoring random forest models.

**Supplementary Figure S5. Construction and performance of the overall mental health scoring random forest models.**

Random forest models fitting 145 survey variables (**Supplementary Table S1**) to the overall mental health (OMH) scoring in the Austria/Tyrol (AT) and Italy/South Tyrol (IT) cohorts were constructed and validated by the 10-fold cross-validation (CV) technique.

(**A**) Top 20 most influential factors contributing to the improvement of to model fit measured as difference in mean squared error ($\Delta$MSE). N numbers of observations are indicated below the plot.

(**B**) Fitted versus true scoring values in the Austria/Tyrol and Italy/South Tyrol cohorts. Spearman's $\rho$ correlation coefficients, means absolute errors (MAE) for the whole-cohort and cross-validation data sets and n numbers of observations are indicated below the plot.

imp.: impaired, pre-cov depr/anxiety: depression or anxiety before COVID-19, tired. day: tiredness at day, prol.: prolonged, SIF: severe illness feeling, #: number, NC: neurocognitive symptoms, GP: general practitioner, persist.: persistent, dim.: diminished, 2Q, 3Q, 4Q: $2^{nd}$, $3^{rd}$, $4^{th}$ quartile, empl.: employment, conc.: concentration, daily medic.: daily medication, # cov in household: number of COVID-19 cases in the household, subj. cov percept.: subjective perception of acute COVID-19, pre-cov sleep disord.: sleep disorder before COVID-19.

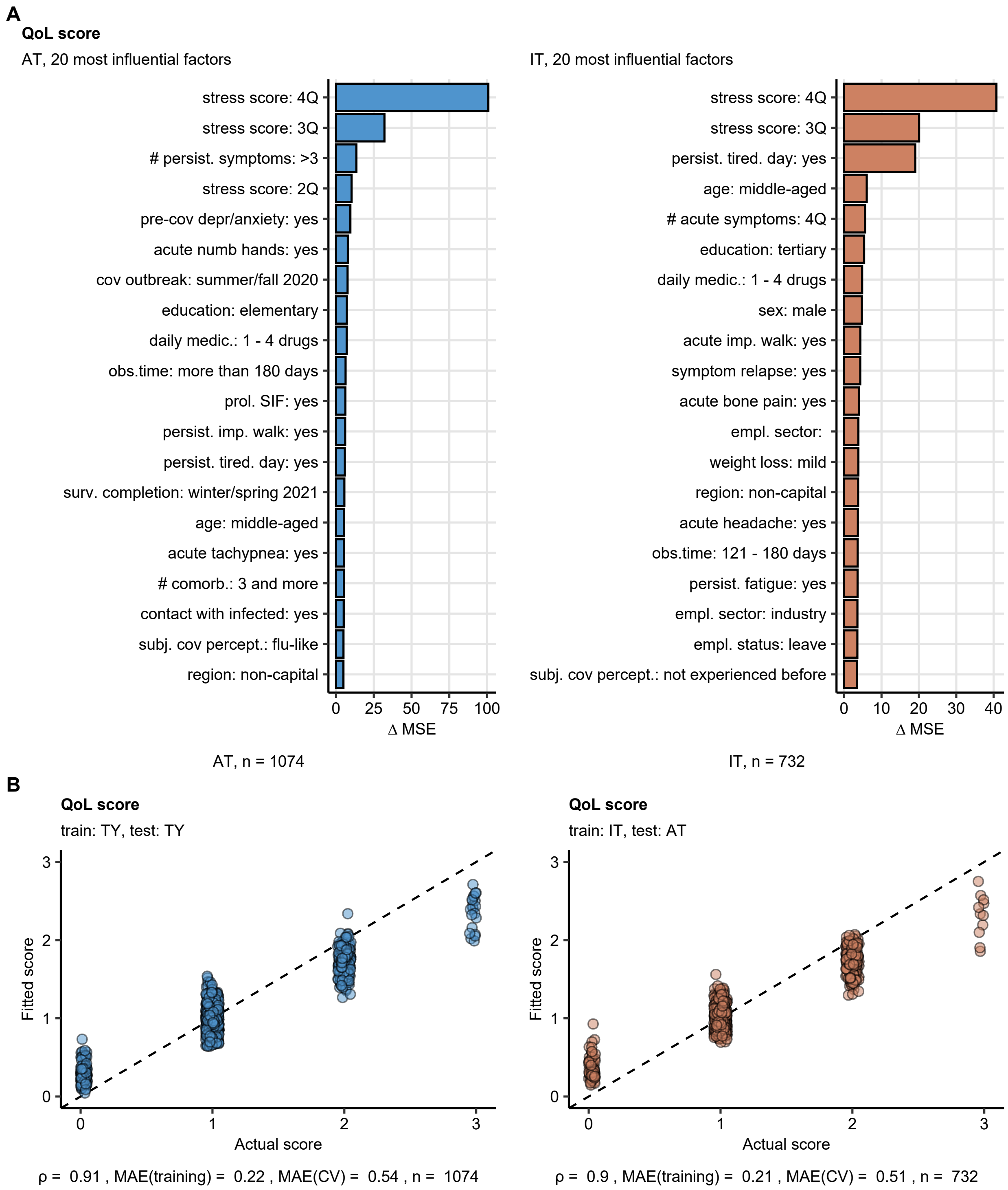

Figure S6: Construction and performance of the quality of life scoring random forest models.

**Supplementary Figure S6. Construction and performance of the quality of life scoring random forest models.**

Random forest models fitting 145 survey variables (**Supplementary Table S1**) to the quality of life (QoL) scoring in the austria/Tyrol (AT) and Italy/South Tyrol (IT) cohorts were constructed and validated by the 10-fold cross-validation (CV) technique.

(**A**) Top 20 most influential factors contributing to the improvement of to model fit measured as difference in mean squared error (ΔMSE). N numbers of observations are indicated below the plot.

(**B**) Fitted versus true scoring values in the Austria/Tyrol and Italy/South Tyrol cohorts. Spearman's $\rho$ correlation coefficients, means absolute errors (MAE) for the whole-cohort and cross-validation data sets and n numbers of observations are indicated below the plot.

imp.: impaired, pre-cov depr/anxiety: depression or anxiety before COVID-19, tired. day: tiredness at day, prol.: prolonged, SIF: severe illness feeling, #: number, NC: neurocognitive symptoms, GP: general practitioner, persist.: persistent, dim.: diminished, 2Q, 3Q, 4Q: $2^{nd}$, $3^{rd}$, $4^{th}$ quartile, empl.: employment, conc.: concentration, daily medic.: daily medication, # cov in household: number of COVID-19 cases in the household, subj. cov percept.: subjective perception of acute COVID-19, pre-cov sleep disord.: sleep disorder before COVID-19.

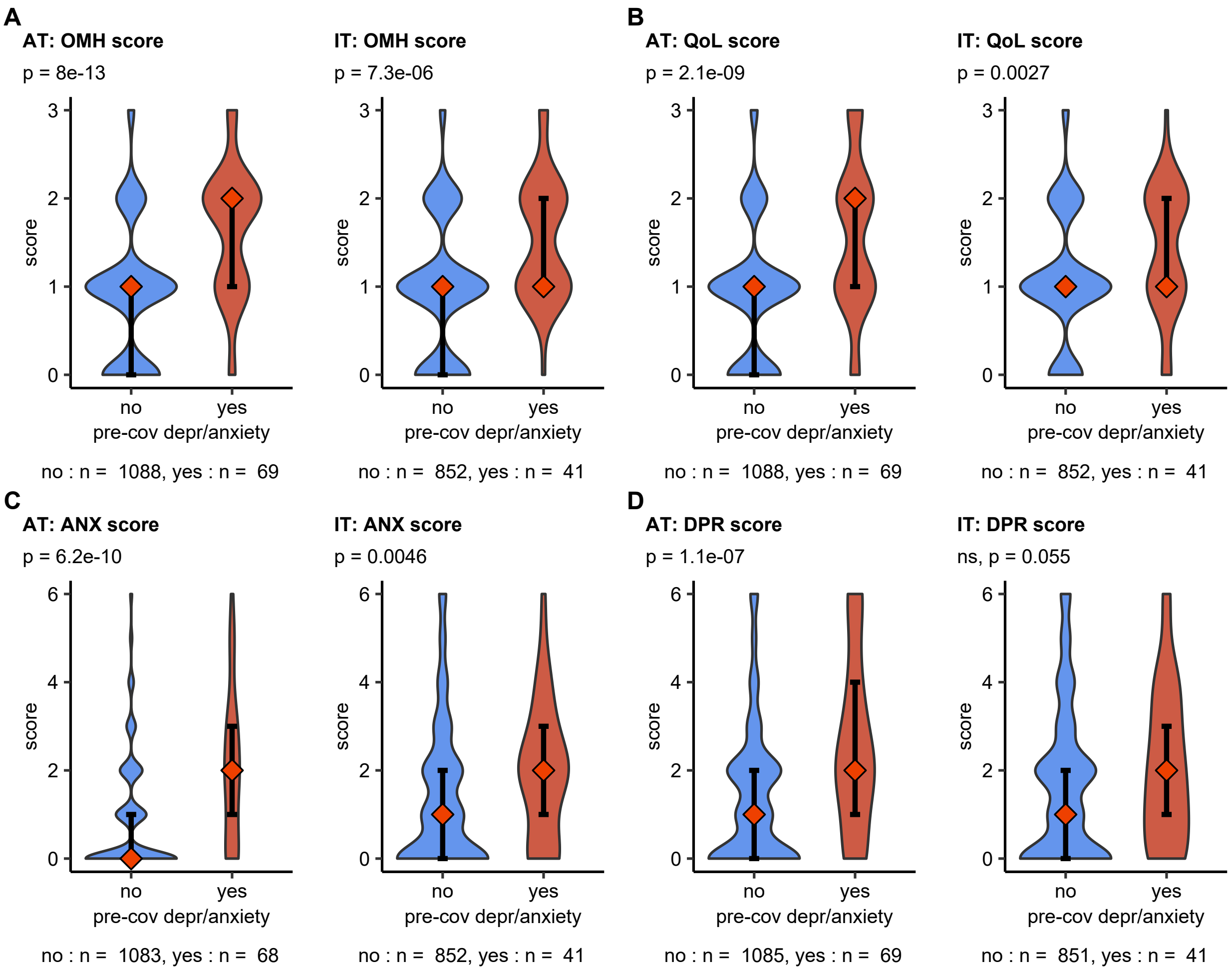

Figure S7: Depression or anxiety before COVID-19 and mental health scoring.

**Supplementary Figure S7. Depression or anxiety before COVID-19 and mental health scoring.**

Association of depression or anxiety before COVID-19 with overall mental health (OMH) (**A**), quality of life (QoL) (**B**), anxiety (ANX) (**C**) and depression (DPR) (**D**) scoring assessed by Mann-Whitney U test. The scoring is presented as violin plots, diamonds with whiskers represent medians with IQRs. P values corrected for multiple comparisons with Benjamini-Hochberg method are shown in plot sub-headings. N numbers of observations are indicated below the plot.

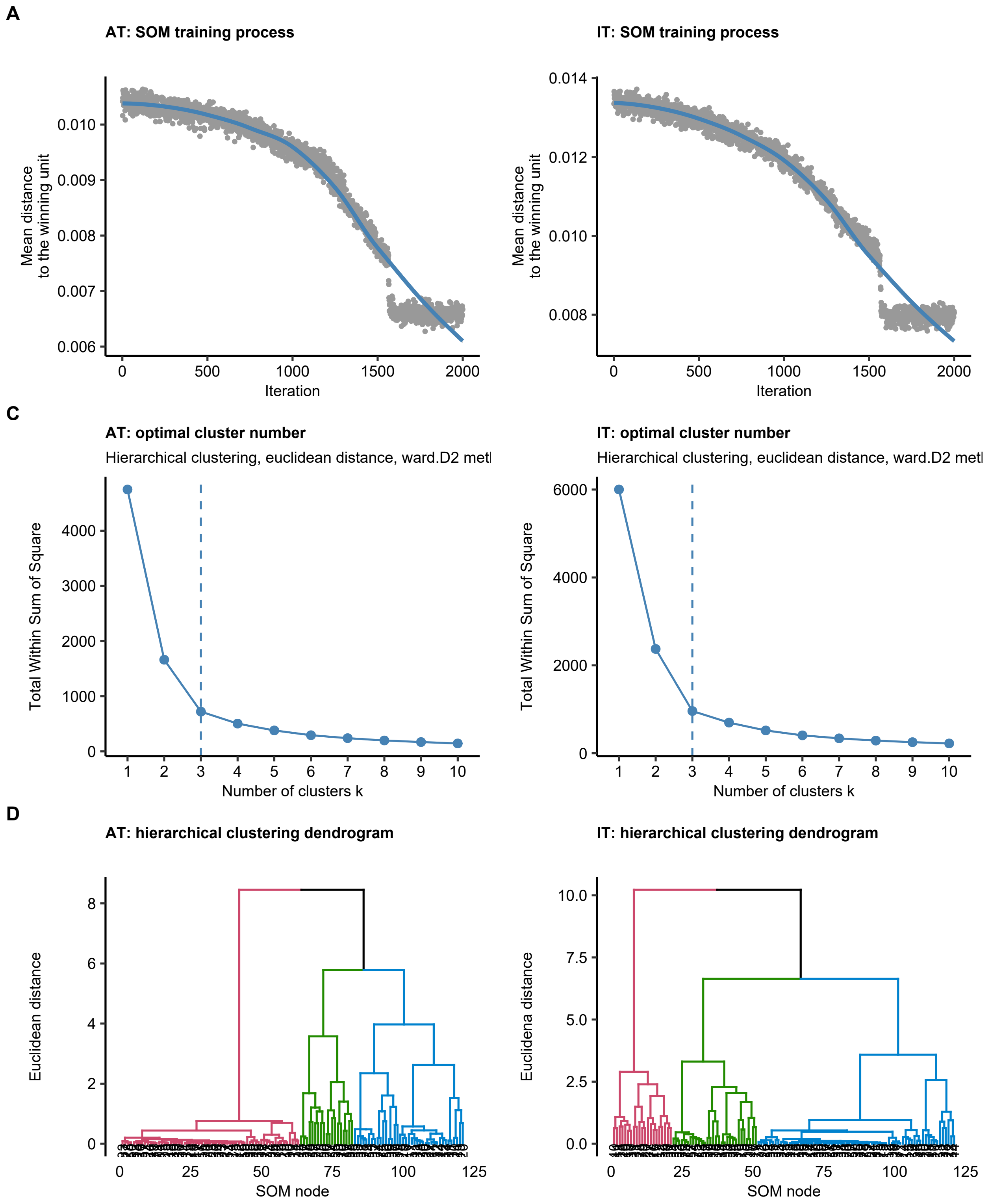

Figure S8: Development of the mental disorder risk clusters.

**Supplementary Figure S8. Development of the mental disorder risk clusters.**

Study participants were assigned to the Low Risk (LR), Intermediate Risk (IR) and High Risk (HR) subsets by clustering analysis of the most influential factors impacting the combined mental health and quality of life scoring (**Figure 2**) with the self-organizing map (SOM, 11 × 11 hexagonal grid, Jaccard distance between participants) and the hierarchical clustering (Ward D2 method, Euclidean distance between the SOM nodes) algorithms as presented in **Figure 4**.

**(A)** Progress of the SOM training procedure visualized as the drop of the mean distance to the winning unit with the algorithm iterations.

**(B)** Determination of the optimal cluster number in hierarchical clustering of the SOM nodes by finding the bend of the total within sum of square curve.

**(C)** Assignment of the SOM nodes to the clusters defined by hierarchical clustering presented in dendrograms.

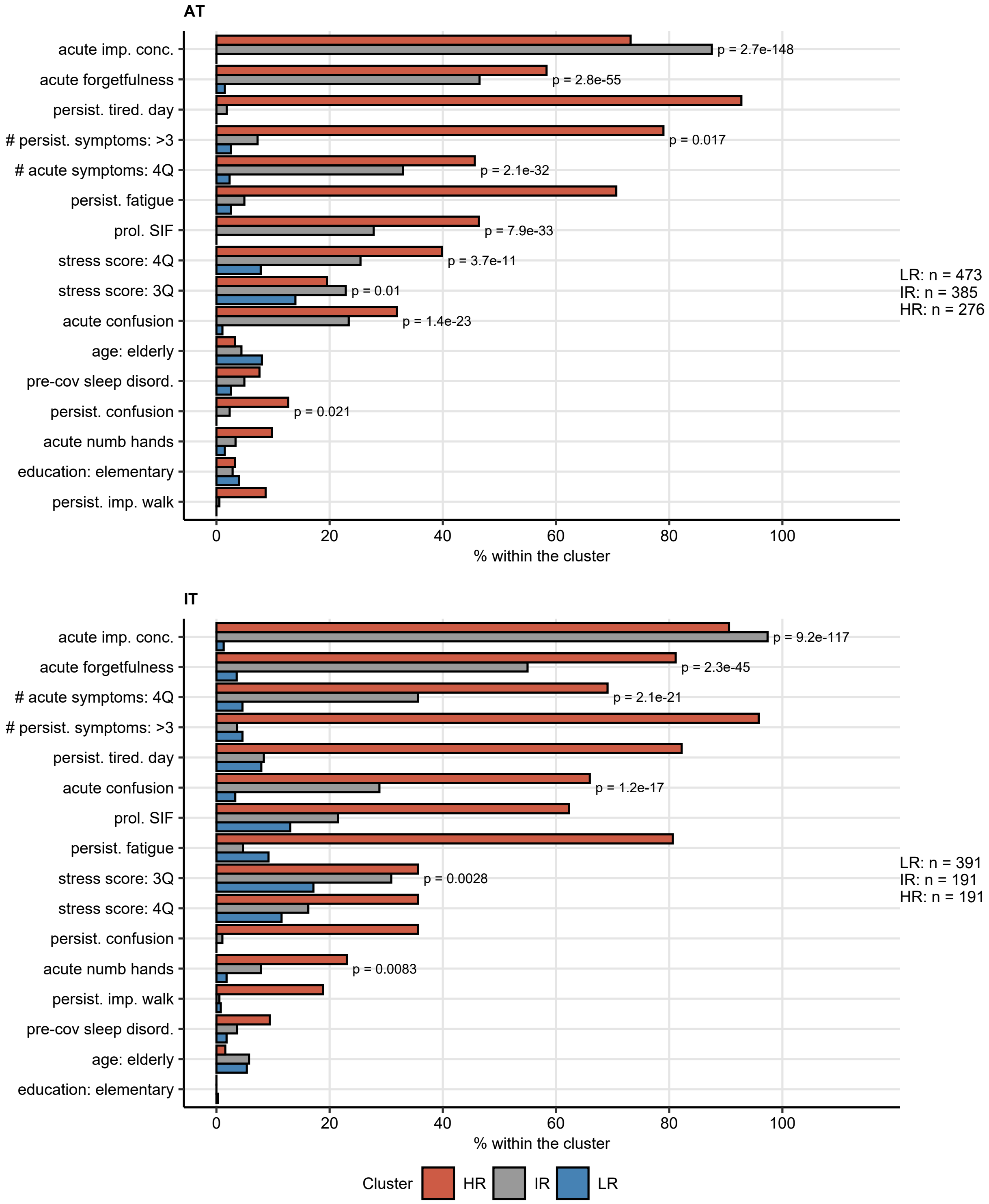

Figure S9: Frequency of the clustering features in the mental disorder risk clusters.

**Supplementary Figure S9. Frequency of the clustering features in the mental disorder risk clusters.**

Study participants were assigned to the Low Risk (LR), Intermediate Risk (IR) and High Risk (HR) subsets as presented in **Figure 4**. Differences in frequency of these features between the Low risk (LR), Intermediate Risk (IR) and High Risk (HR) clusters were assessed by $\chi^2$ test. P values corrected for multiple comparisons with Benjamini-Hochberg method are presented for the significant comparisons. N numbers of individuals assigned to the clusters are presented next to the plots.

prol.: prolonged, SIF: severe illness feeling, imp.: impaired, conc.: concentration, #: number, tired.day.: tiredness at day, pre-cov sleep disord.: sleep disorder before COVID-19, 3Q, 4Q: 3$^{rd}$ and 4$^{th}$ quartile, persist.: persistent.

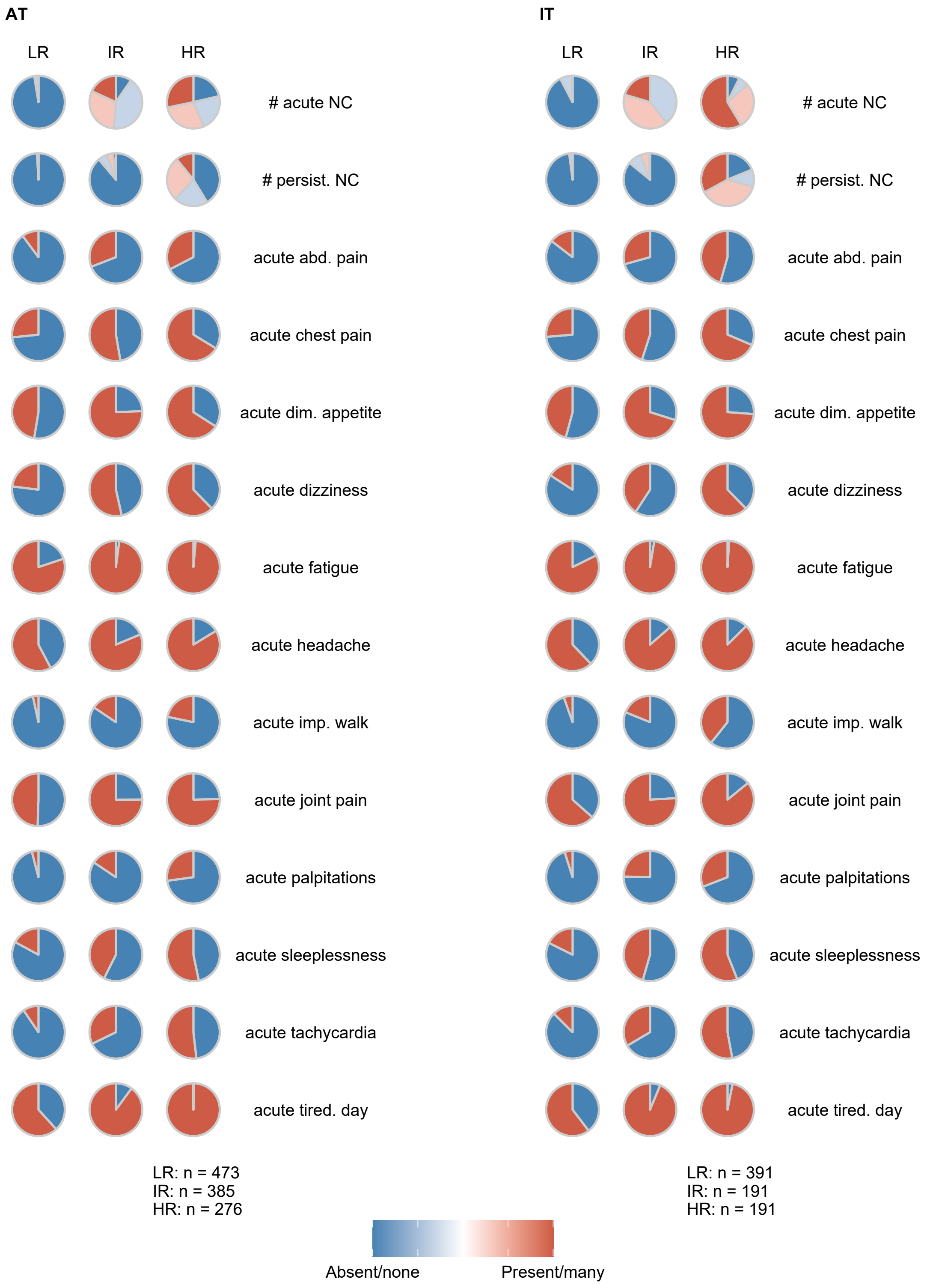

Figure S10: Frequency of the most significant differing features in the mental disorder risk clusters.

**Supplementary Figure S10. Frequency of the most significant differing features in the mental disorder risk clusters.**

Study participants were assigned to the Low Risk (LR), Intermediate Risk (IR) and High Risk (HR) subsets as presented in **Figure 4**. Differences in frequency of 130 survey variables not used for the cluster definition (**Figure 4**, **Supplementary Table S1**) between the risk clusters were compared by $\chi^2$ test. P values were corrected for multiple comparisons with Benjamini-Hochberg method. Frequencies of the most significant features within the mental disorder risk clusters are shown. N numbers of individuals assigned to the clusters are presented next to the plots.

NC: neurocognitive symptoms, #: number, persist.: persistent, abd.: abdominal, dim.: diminished, subj. cov percept.: subjective perception of acute COVID-19.

### References

1. Phenotyping of acute and persistent COVID-19 features in the outpatient setting: exploratory analysis of an international cross-sectional online survey. *medRxiv.* Published online August 2021:2021.08.05.21261677. doi:10.1101/2021.08.05.21261677

2. Löwe B, Wahl I, Rose M, et al. A 4-item measure of depression and anxiety: Validation and standardization of the Patient Health Questionnaire-4 (PHQ-4) in the general population. *Journal of Affective Disorders.* 2010;122(1-2):86-95. doi:10.1016/j.jad.2009.06.019

3. Löwe B, Spitzer RL, Zipfel S, Herzog W. *Auflage Manual 17.07.* Vol 9.; 2002:44-45.

4. Beutel TF, Zwerenz R, Michal M. Psychosocial stress impairs health behavior in patients with mental disorders. *BMC Psychiatry.* 2018;18(1). doi:10.1186/s12888-018-1956-8

5. Gräfe K, Zipfel S, Herzog W, Löwe B. Screening psychischer störungen mit dem "Gesundheitsfragebogen für Patienten (PHQ-D)". Ergebnisse der Deutschen validierungsstudie. *Diagnostica.* 2004;50(4):171-181. doi:10.1026/0012-1924.50.4.171

6. Health after COVID-19 in Tyrol. Mental Health after COVID-19 in Tyrol. Accessed September 9, 2021. https://im2-ibk.shinyapps.io/mental_health_dashboard/

7. Wickham H, Averick M, Bryan J, et al. Welcome to the Tidyverse. *Journal of Open Source Software.* 2019;4(43):1686. doi:10.21105/joss.01686

8. Wickham H. *ggplot2: Elegant Graphics for Data Analysis.* 1st ed. Springer-Verlag; 2016. https://ggplot2.tidyverse.org

9. Wilke CO. *Fundamentals of Data Visualization: A Primer on Making Informative and Compelling Figures.* 1st ed. O'Reilly Media; 2019:389.

10. Benjamini Y, Hochberg Y. Controlling the False Discovery Rate: A Practical and Powerful Approach to Multiple Testing. *Journal of the Royal Statistical Society: Series B (Methodological).* 1995;57(1):289-300. doi:10.1111/j.2517-6161.1995.tb02031.x

11. Breiman L. Random forests. *Machine Learning.* 2001;45(1):5-32. doi:10.1023/A:1010933404324

12. Kuhn M. Building predictive models in R using the caret package. *Journal of Statistical Software.* 2008;28(5):1-26. doi:10.18637/jss.v028.i05

13. Strobl C, Boulesteix AL, Kneib T, Augustin T, Zeileis A. Conditional variable importance for random forests. *BMC Bioinformatics.* 2008;9(1):1-11. doi:10.1186/1471-2105-9-307

14. Croux C, Filzmoser P, Oliveira MR. Algorithms for Projection-Pursuit robust principal component analysis. *Chemometrics and Intelligent Laboratory Systems.* 2007;87(2):218-225. doi:10.1016/j.chemolab.2007.01.004

15. Austrian Agency for Health and Food Safety (AGES). Epidemiologische Abklärung Covid 19. Accessed August 5, 2021. https://www.ages.at/themen/krankheitserreger/coronavirus/epidemiologische-abklaerung-covid-19/

16. Istituto Superiore di Sanitá (ISS). Integrated surveillance of COVID-19 in Italy. Accessed August 5, 2021. https://www.epicentro.iss.it/coronavirus/bollettino/Bollettino-sorveglianza-integrata-COVID-19_7-luglio-2021.pdf

17. Borenstein M, Hedges LV, Higgins JPT, Rothstein HR. A basic introduction to fixed-effect and random-effects models for meta-analysis. *Research Synthesis Methods*. 2010;1(2):97-111. doi:10.1002/jrsm.12

18. Balduzzi S, Rücker G, Schwarzer G. How to perform a meta-analysis with R: A practical tutorial. *Evidence-Based Mental Health*. 2019;22(4):153-160. doi:10.1136/ebmental-2019-300117

19. Vesanto J, Alhoniemi E. Clustering of the self-organizing map. *IEEE Transactions on Neural Networks*. 2000;11(3):586-600. doi:10.1109/72.846731

20. Kohonen T. *Self-Organizing Maps*. Vol 30. Springer Berlin Heidelberg; 1995. doi:10.1007/978-3-642-97610-0

21. Vesanto J, Vesanto J, Himberg J, Alhoniemi E, Parhankangas J. Self-organizing map in Matlab: the SOM toolbox. *IN PROCEEDINGS OF THE MATLAB DSP CONFERENCE*. Published online 1999:35—-40. http://citeseerx.ist.psu.edu/viewdoc/summary?doi=10.1.1.97.179
